# Multi-modal sleep staging in the clinic for REM sleep behaviour disorder

**DOI:** 10.64898/2026.06.30.26356905

**Authors:** Katarina M. Gunter, Nivedita Bijlani, Gary Dennis, Christine Lo, Timothy Quinnell, Mkael Symmonds, Jessica Welch, Pietro-Luca Ratti, Michele T. Hu, Mauricio Villaroel

## Abstract

Accurate REM identification is critical for diagnosing REM sleep behaviour disorder (RBD), yet many automated sleep-staging systems, especially single-channel EEG models trained on healthy cohorts, do not generalise well to real-life polysomnography (PSG) performed in patients. We compared a feature-based Random Forest (RF) model tuned for RBD with a state-of-the-art single- EEG deep architecture (AttnSleep), and assessed the impact of cohort adaptation and multimodal inputs (EEG, EOG, EMG, ECG). Experiments used 89 multi-site in-clinic PSGs (SleepWearables Phase-1) plus 53 MASS healthy controls (mean age 63 *±* 5 years), with 10-fold cross-validation and out-of-fold evaluation. When applied out-of-the-box after training on open-source healthy datasets, the RF outperformed AttnSleep on the clinical cohort (Cohen’s *κ* = 0.56 versus 0.49), with the largest difference in REM sleep in RBD subjects (RF Cohen’s *κ* = 0.51 versus AttnSleep Cohen’s *κ* = 0.18), highlighting limited cross-cohort generalisation. Cohort-specific retraining substantially improved AttnSleep, and the multi-modal model further improved overall agreement (Cohen’s *κ* 0.61 versus 0.63), with the largest stage-level gains in N1 and N3; in RBD subjects, however, overall agreement was unchanged (Cohen’s *κ* 0.46 for both configurations). Attention-based modality analysis identified EEG as the dominant signal, increased EOG contribution during REM and N1, and elevated ECG importance during N3. In RBD subjects, EOG weighting increased relative to subjects without RBD (Δ = +0.096) while EEG weighting fell (Δ = *−*0.056), suggesting physiological relevance. Guided by these weights, a reduced four-channel EEG model matched full multimodal performance in non-RBD subjects, and adding EOG matched it overall (Cohen’s *κ* = 0.63) while achieving the best performance in RBD (Cohen’s *κ* = 0.49) and improved REM classification in RBD (60.6% compared to 53.0% recall). Inclusion of EOG also halved inter-dataset variability in REM staging between the clinical and MASS cohorts. Nonetheless, staging performance in RBD remained lower than in controls, particularly for REM, and within the RBD subgroup only the REM improvement from the four-channel EEG + EOG configuration reached statistical significance at this sample size (*n* = 22). These results highlight (1) the limited generalisability of minimal-sensor models trained on healthy cohorts, (2) the value of mixed cohort-specific training, and (3) that the composition of the sensor set matters more than its size: targeted addition of EOG improved REM staging in RBD specifically, whereas full multimodal integration improved performance mainly outside the RBD subgroup.

## 1 Introduction

REM sleep behaviour disorder (RBD) is a parasomnia defined by the loss of normal muscle atonia during REM sleep, referred to as REM sleep without atonia (RSWA). It is clinically characterised by dream enactment behaviours including vocalisations, movements of the body, upper and lower limbs [1, 2]. RBD has a prevalence of approximately 0.5–1%. However, prevalence rises substantially with age and is likely under-reported [3, 4]. It predominantly affects older adults, more commonly males over the age of 50, and is strongly associated with neurodegenerative disease. Longitudinal evidence suggests that 70–80% of individuals with idiopathic RBD will develop an *α*-synucleinopathy, such as Parkinson’s disease (PD), dementia with Lewy bodies (DLB), or multiple system atrophy (MSA) - with approximately 75% converting within 12 years [5].

RBD is also emerging as a condition of specific concern due to its association with contact sport participation and long-term neurodegenerative risk. Adams et al. [6] found that in a cohort of 247 deceased male contact sport athletes with neuropathologically confirmed chronic traumatic encephalopathy (CTE), 32% had experienced probable RBD during life, a prevalence substantially higher than the 1% observed in the general population. The odds of RBD symptoms increased by approximately 4% for each additional year of contact sport participation [6]. REM sleep architecture is especially vulnerable to disruption from intense athletic effort. Roig-Uribe et al. [7] examined concussion history in 199 polysomnography (PSG) confirmed iRBD patients and 168 matched controls. Prior concussion was significantly more common in iRBD patients than controls (21.1% vs 10.1%, p = 0.004), with a mean interval of 43 years between concussion and iRBD diagnosis. These findings also extend to living retired athletes [8].

Detection of RBD remains challenging. Many patients are unaware of their symptoms, which may only be reported by bed partners. Confirming a diagnosis requires PSG to demonstrate both RSWA and dream enactment. Accurate identification of REM sleep is therefore critical, yet sleep staging in RBD patients is particularly challenging due to their abnormal REM physiology and behaviour. Manual sleep staging is highly resource-intensive, demanding specialist expertise and is time-consuming. In the UK, patients often face waiting times of up to 12 months for a sleep clinic appointment [9]. As most individuals with RBD go on to develop an *α*-synucleinopathy, timely and scalable assessment is clinically important for monitoring and for evaluating the effectiveness of disease-modifying therapies.

Automatic sleep staging has evolved from the selection of hand-crafted features with classical machine learning to end-to-end deep learning methods, with an increasing emphasis on minimal-sensor configurations (e.g., single-channel EEG) [10, 11, 12, 13, 14]. While large public datasets (Sleep-EDF [15], MASS [16], PhysioNet [17], SHHS [18]) have enabled standardised benchmarking, most validation focuses on relatively homogeneous or community cohorts. External validation in diverse clinical populations remains uncertain; notable exceptions include YASA [10], trained across seven open-source datasets including subjects with suspected sleep-disordered breathing.

RBD poses specific challenges for automated staging that limit direct translation of models trained on non-RBD cohorts: (i) RSWA violates the standard REM-atonia assumption, so EMG-based features can misclassify REM as N1 or wake; (ii) phasic muscle bursts and dream-enactment movements introduce artefacts that degrade EEG/EOG features; (iii) REM fragmentation with frequent arousals blurs stage boundaries; and (iv) class imbalance and scorer disagreement around REM. Consistent with this, a random-forest (RF) model trained on hand-crafted EEG/EOG/EMG features achieved lower agreement in RBD than in controls, particularly for REM [19], with performance below typical inter-rater variability between human scorers [20, 21, 22]. More broadly, model performance is known to vary with dataset characteristics, including sleep disorder mix and demographics [23, 24]. Whilst existing work suggests reduced REM agreement in RBD compared with controls [19], there has been no rigorous, head-to-head evaluation of classical feature-engineered approaches explicitly tailored to RBD physiology against modern end-to-end deep learning architectures in real-world, multi-site clinical PSGs. Moreover, there lacks evidence regarding the contribution of individual modalities (EEG, EOG, EMG, ECG) to staging performance in RBD, limiting decisions about sensor reduction versus clinical fidelity.

In this work, we directly compare an established classical Random Forest model tailored to RBD/controls and built on hand-crafted multimodal features [19] against a state-of-the-art, single-EEG deep model [25]. We evaluate whether the latter’s complexity and learned features actually generalise to real-life, in-clinic, multi-site PSGs - especially in RBD. We then assess the added value of moving beyond a minimal sensor set-up by re-introducing the full clinical montage (EEG, EOG, EMG, ECG) into the deep architecture, and quantify gains from cohort-specific model training. Our evaluation focuses on clinically relevant endpoints: stage-wise performance with emphasis on REM, robustness across sites, and the importance of each modality in sleep staging across the mixed cohort. Our contributions are as follows:

- Benchmarking an RBD-focused Random Forest (RF) model and a modern deep learning model on multi-site in-clinic PSG data.
- Cohort adaptation: re-training the deep learning model on our clinical cohort and assess the performance.
- We extend the deep learning architecture to incorporate multiple sensing modalities (EEG, EOG, EMG, ECG), and evaluate both overall performance gains and modality-specific contributions.
- Clinically informed modality reduction: leveraging modality importance derived from the multi-modal model, we develop a simplified architecture using only the most informative signals, evaluating whether diagnostic performance can be maintained while reducing sensing burden.
- We perform subgroup analyses with a focus on REM staging in RBD, highlighting where the model succeeds and where it fails in capturing disease-specific sleep dynamics.

## 2 Methods

### 2.1 Datasets

In this work we use four datasets: Sleep-EDF-20 (and expanded Sleep-EDF-78 counterpart) [15], the Sleep Heart Health Study (SHHS) [18], the Montreal archive of sleep studies cohort 1, subset 1 (MASS) [16], and the SleepWearables study. Sleep-EDF-20, Sleep-EDF-78, and SHHS are open-source PSG datasets commonly used for automated sleep staging research and were used to train the baseline AttnSleep model. Models were evaluated on the MASS and SleepWearables dataset combined.

An overview of the demographics of subjects in each dataset are given in table 1. The instrumentation along with recording set-up and associated sampling frequencies for each dataset are shown in table 2.

**Table 1:** Overview of the subjects and demographics of the datasets used for analysis. *µ*: mean. SD: standard deviation.

| Dataset | Site | Diagnosis group | N Subjects | Age ($\mu \pm SD$) | Sex (% male) |
| --- | --- | --- | --- | --- | --- |
| SleepWearables (SW) | John Radcliffe | HC | 13 | 69.8 (16.2) | 61.5 |
|  |  | RBD | 15 | 67.7 (5.2) | 86.7 |
|  |  | Other | 14 | 48.6 (19.2) | 71.4 |
|  | Sheffield | HC | 0 | - | - |
|  |  | RBD | 7 | 61.3 (6.5) | 100.0 |
|  |  | Other | 18 | 40.4 (19.9) | 50.0 |
|  | Papworth | HC | 6 | 63.8 (20.5) | 50.0 |
|  |  | RBD | 10 | 61.1 (6.2) | 80.0 |
|  |  | Other | 10 | 50.5 (16.1) | 40.0 |
|  | Total SW | HC | 19 | 67.9 (17.3) | 57.9 |
|  |  | RBD | 32 | 64.2 (6.5) | 87.5 |
|  |  | Other | 42 | 45.5 (19.0) | 54.8 |
| MASS SS1 | - | Elderly HC | 53 | 63.0 (5.0) | 64.2 |
| Sleep-EDF-20 | - | HC | 20 | 28.7 (2.9) | 50.0 |
| Sleep-EDF-78 | - | HC | 78 | 58.8 (22.2) | 47.4 |
| SHHS | - | HC | 329 | 55.6 (10.2) | 18.8 |

**Table 2:** Summary of the physiological signals and in-clinic PSG channels available for Phase 1 of the SleepWearables study and open-source control datasets (MASS, Sleep-EDF-78, and SHHS). The sampling frequency is given as the minimum sampling frequency present in the data from a given site. Sleep-EDF-20 has the same specifications as the expanded Sleep-EDF-78.

| Dataset | Site | Device | Signal | Description | Sampling frequency (Hz) |
| --- | --- | --- | --- | --- | --- |
| SleepWearables | JR | Natus EMU40/<br>Natus Trex | EEG | 20-channel montage | 200 |
|  |  |  | EOG | 2-channel (left & right) | 200 |
|  |  |  | EMG | 2 chin, 2 bilateral anterior tibialis | 200 |
|  |  |  | ECG | 2-lead | 200 |
|  |  |  | Pulse Rate | Pulse oximeter | 200 |
|  |  |  | SpO2 | Pulse oximeter | 200 |
|  |  |  | Air Flow | Nasal cannula & thermistor | 200 |
|  |  |  | Respiratory Effort | Abdominal and thoracic plethysmography belt | 200 |
|  |  |  | Body Position | Derived from 3-axis accelerometer | 200 |
| SleepWearables | Sheffield | Natus EMU40/<br>Natus Trex | EEG | 20-channel montage | 256 |
|  |  |  | EOG | 2-channel (left & right) | 256 |
|  |  |  | EMG | 2 chin, 2 bilateral anterior tibialis | 256 |
|  |  |  | ECG | 2-lead | 256 |
|  |  |  | Pulse Rate | Pulse oximeter | 256 |
|  |  |  | SpO2 | Pulse oximeter | 256 |
|  |  |  | Air Flow | Nasal cannula | 256 |
|  |  |  | Respiratory Effort | Abdominal and thoracic plethysmography belt | 256 |
|  |  |  | Body Position | Derived from 3-axis accelerometer | 256 |
| SleepWearables | Papworth | Natus Embla | EEG | 2 central & 2 occipital | 200 |
|  |  |  | EOG | 2-channel (left & right) | 200 |
|  |  |  | EMG | 3 chin, 2 bilateral anterior tibialis | 200 |
|  |  |  | Pulse Rate | Pulse oximeter | 3 |
|  |  |  | SpO2 | Pulse oximeter | 3 |
|  |  |  | Air Flow | Nasal cannula & thermistor | 20 |
|  |  |  | Respiratory Effort | Abdominal and thoracic plethysmography belt | 10 |
|  |  |  | Body Position | Derived from 3-axis accelerometer | 10 |
| MASS SS1 | - | Grass<br>Model 15 | EEG | 17 or 19 channel | 256 |
|  |  |  | EOG | 2-channel (left & right) | 256 |
|  |  |  | EMG | 3 chin, 2 bilateral anterior tibialis | 256 |
|  |  |  | ECG | single-lead (D-I) | 512 |
|  |  |  | Pulse Rate | Pulse oximeter | 256 |
|  |  |  | SpO2 | Pulse oximeter | 256 |
|  |  |  | Air Flow | Nasal cannula & thermistor | 32 & 128 |
| | | | Respiratory Effort | Abdominal ± thoracic plethysmography belt | 128 |
| Sleep-EDF-78 / Sleep-EDF-20 | - | - | EEG | Fpz-Cz and Pz-Oz | 100 |
|  |  |  | EOG | 2-channel (left & right) | 100 |
|  |  |  | EMG | Submental chin EMG envelope | 1 |
| SHHS | - | Compumedics<br>P-series | EEG | C3-A2 and C4-A1 | 125 |
|  |  |  | EOG | 2-channel (left & right) | 50 |
|  |  |  | EMG | 3 chin, 2 bilateral anterior tibialis | 125 |
|  |  |  | ECG | bipolar single-channel | 125 |
|  |  |  | Pulse Rate | Pulse oximeter | 1 |
|  |  |  | SpO2 | Pulse oximeter | 1 |
|  |  |  | Air Flow | thermistor | 10 |
| | | | Respiratory Effort | Abdominal ± thoracic plethysmography belt | 10 |

#### 2.1.1 Sleep-EDF-20/78

Sleep-EDF-20 is a subset of the Sleep-EDF Expanded dataset and includes overnight PSG recordings from 20 healthy caucasian subjects in the age range of 21-35 years [15]. Subjects were not on medication. Recordings contain EOG and two EEG channels (Fpz-Cz and Pz-OZ. The recordings were annotated according to standard sleep staging guidelines (30-second epochs following Rechtschaffen & Kales standards). It is widely used as a benchmark dataset for sleep stage classification. Sleep-EDF-78 is a larger subset of the same Sleep-EDF Expanded database, containing recordings from 78 subjects. It provides a more diverse sample across multiple nights and subjects.

#### 2.1.2 SHHS

SHHS is a large-scale, community-based cohort study containing thousands of overnight PSG recordings [18]. The dataset includes multi-channel physiological signals (including EEG, EOG, EMG, ECG, respiratory signals, oxygen saturation) along with sleep stage annotations (following Rechtschaffen & Kales standards). Here we use the SHHS visit 1 cohort, which has a fixed sampling rate of 125 Hz. This cohort contains recordings from over 6,441 individuals, with a minimum age of 40 years. In this work we use a subset of the cohort (329), filtered based on sleep-related and cardiovascular diseases, as described in the AttnSleep publication [25].

#### 2.1.3 SleepWearables study

The primary aim of the SleepWearables study was to assess the ability of a photoplethysmogram (PPG) sensor, actigraph (motion sensor worn at the wrist) and portable sleep monitoring device to detect RBD compared to the gold standard PSG and screening (RBDSQ) tests. Secondary aims were to use these devices to (i) assess RBD in a Parkinson’s disease cohort versus controls and participants with confirmed RBD, and (ii) track longitudinal progression in RBD and PD, including response to treatment. This study was a collaboration between the Nuffield Department of Clinical Neurosciences (NDCN) and the University of Oxford, funded by the Oxford Biomedical Research Centre (BRC). It was approved by the South Central – Oxford Research Ethics Committee with the reference number 17/SC/0631.

The first phase of the study included healthy controls and participants referred for PSG for the investigation of suspected sleep disorders at three sites in the UK: the John Radcliffe (JR) hospital (Oxford), the Royal Papworth hospital (Cambridge), and Royal Hallamshire hospital (Sheffield). Data was collected for one night in the sleep clinic, on site. The participants wore the in-house gold standard PSG system, as well as the remote monitoring PSG device.

The criteria for inclusion were that subjects were aged 18 or over, and did not exhibit any cognitive impairment which may impact their ability to provide informed consent. The control participants had no diagnosis of RBD or PD and no self-reported sleep problems, confirmed via sleep screening questionnaires.

Subjects wore an in-clinic gold standard PSG kit. Sleep stage annotations were given in 30 second epochs according to AASM criteria. Data were collected in all subjects for one night, unless otherwise specified. As the “patient” population were referred to the clinic for sleep disturbances, this serves as their diagnostic PSG. The diagnosis was provided by a trained sleep clinician. Table 1 summarises the PSG dataset by site, specifying the number of healthy controls (HC), RBD diagnosed subjects, and a group termed “other” - subjects without RBD, but other sleep disorders. The diagnoses within the “other” group include: sleep apnoea or obstructive sleep apnoea, insomnia, narcolepsy, periodic leg movements (PLMs), periodic leg movement disorder (PLMD), and idiopathic hypersomnia. Two subjects were without a diagnosis group, and one subject had two nights of PSG recordings.

#### 2.1.4 MASS

MASS cohort 1, subset 1 [16] is a publicly available polysomnography (PSG) dataset comprising overnight recordings from healthy young adults. The dataset includes multi-channel EEG along with standard PSG signals such as EOG and EMG, and provides expert-scored sleep stage annotations in 30-second epochs according to standardized sleep staging criteria (AASM). Cohort 1, subset 1 is comprised of recordings from elderly healthy controls, and was included in this work as an additional control dataset.

## 3 Models

A feature-based multi-modal Random Forest (RF) model [19] was compared to a “state-of-the-art” deep learning model (AttnSleep, [25]), which uses one EEG channel to classify sleep stages in healthy controls. We refer to these as baseline models. We then modified the AttnSleep model to use all available EEG, as well as EOG, EMG and ECG as inputs - here termed the multi-modal model.

### 3.1 Baseline models

Two baseline approaches were evaluated: a feature-based Random Forest (RF) model [19] and the deep learning model AttnSleep [25].

The RF model uses three PSG channels (EEG: C4-A1 or C3-A2, EOG: ROC–LOC difference, and chin EMG). Following preprocessing (described in section 4.2), 156 handcrafted features were extracted from 10-second windows, including standard time- and frequency-domain descriptors such as spectral power in canonical EEG bands, zero-crossing rate, and the EMG atonia index. Features were averaged over 30-second epochs and used as input to the RF classifier. The full list of features are given in the supplementary material table 17.

AttnSleep operates directly on raw EEG. Based upon a 30-second EEG input signal (the C4-A1 or Fpz-Cz channel), high frequency and low frequency features are extracted using a smaller kernel and wide kernel CNN, respectively. AttnSleep attempts to highlight the most important features using adaptive feature recalibration. This is followed by a temporal context encoder which models the temporal dependencies in the input signal using multi-head attention. The final layer is a classification layer, which uses a class-aware loss function. The model was originally designed for single-channel EEG sleep staging in healthy control datasets.

In this work, AttnSleep serves as the deep learning baseline, against which the modified multi-modal configurations are compared.

### 3.2 Multi-modal model

The multi-modal model is built upon the original AttnSleep architecture, with each modality being processed by a multi-resolution convolutional neural network (MRCNN). The MRCNN blocks are designed to extract features at multiple frequency scales, based on what is physiologically appropriate for that signal. For each modality, we follow the original architecture by including two branches in the MRCNN to capture high and low frequency information, respectively. An outline of the modified architecture is shown in fig. 1.

**Figure 1:**
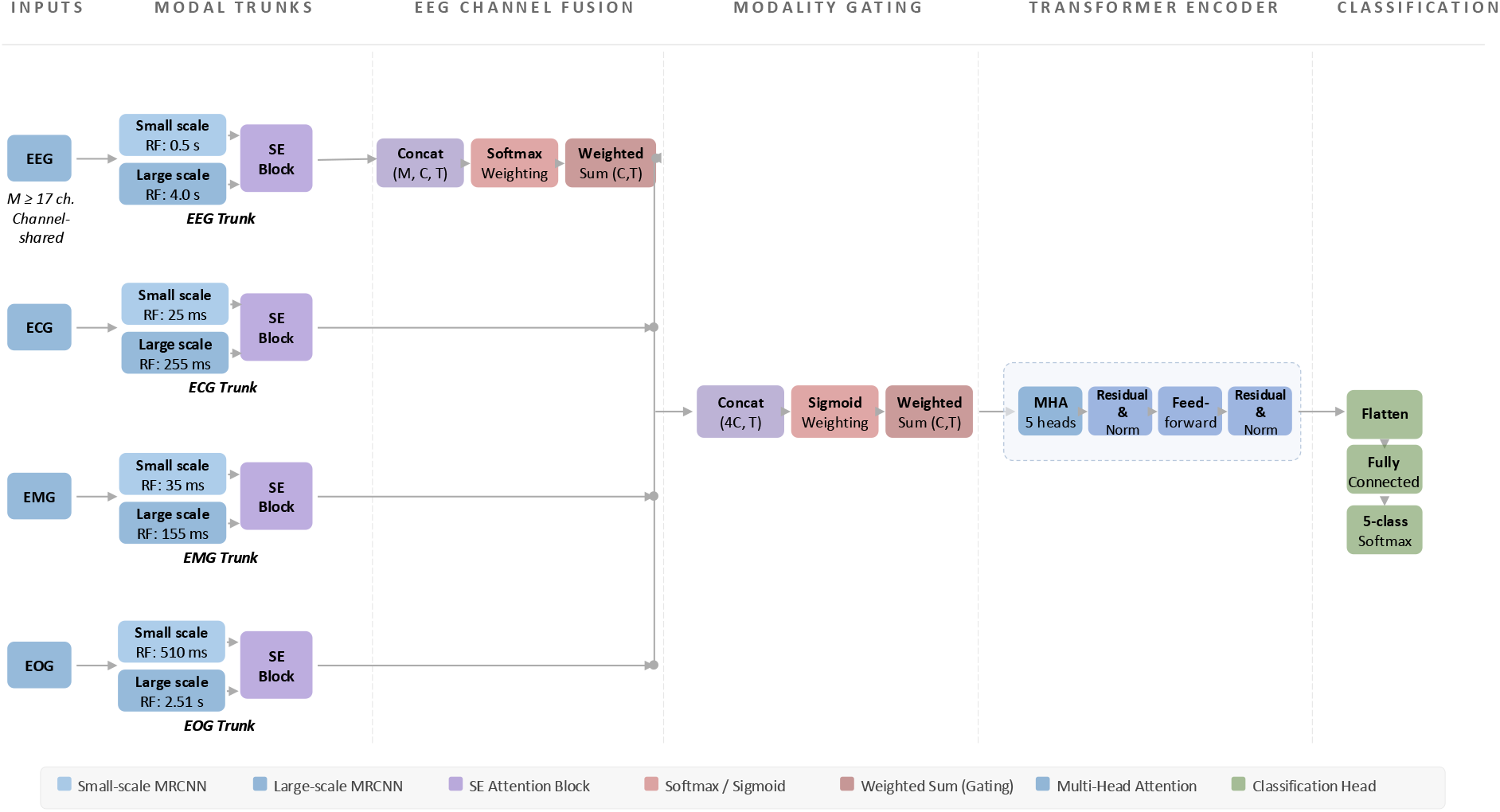
Modified multi-modal AttnSleep feature extraction architecture. Final output features (M: number of EEG channels, C: number of feature maps (30), and T: temporal size (80)) are sent to the temporal context encoder and classification layer from the original architecure.

EEG branch: For EEG, multiple electrode channels are provided simultaneously. Each channel is passed independently through the shared MRCNN from the original AttnSleep model, where the kernel sizes are k=50 and k=400. The per-channel feature maps are subsequently aggregated by a learned channel-fusion module. For each EEG channel, a small two-layer multi-layer perceptron (MLP) produces a scalar importance weight. A softmax across channels ensures that only channels marked as valid by the input mask contribute meaningfully to sleep stage prediction, while noisy or missing channels are suppressed. The weighted sum produces a single fused EEG feature representation of shape (*C, T*), where *C* is the number of feature maps (30), and *T* is the temporal dimension (80).

ECG branch: The ECG branch consists of two parallel convolutional paths designed to capture both fine-scale morphology (e.g., QRS complexes) and broader cardiac cycle dynamics. A small-kernel branch (kernel size 5, receptive field 70 ms) captures high-frequency QRS morphology, while a medium-kernel branch (kernel size 51, RF 460 ms) captures cardiac cycle dynamics. The outputs are concatenated, passed through a squeeze and excitation (SE) residual block to recalibrate channel activations, and adaptively pooled to the temporal dimension *T*.

EMG branch: EMG requires sensitivity to both transient phasic bursts and tonic background muscle tone. The small branch (kernel size 7) uses short kernels with gentle pooling, ensuring preservation of high-frequency bursts characteristic of phasic EMG. The large branch (kernel size 31) begins with a wider kernel and stronger downsampling, capturing lower frequency changes associated with muscle tone or atonia. Both branches are then concatenated, passed through a dropout layer, and recalibrated via an SE residual block before temporal alignment via adaptive pooling.

EOG branch: EOG signals encode both rapid eye movements (100–500 ms) and slower rolling eye movements in N1 sleep. To accommodate these scales, the EOG trunk also employs two convolutional branches. The small branch (kernel size 51) captures high frequency saccadic events, while the large branch (kernel size 251) uses an extended kernel to capture slow eye movements. The two outputs are concatenated, recalibrated by an SE residual block, and temporally normalised.

Masking and modality dropout: For non-EEG modalities, missing inputs are handled by zeroing out the corresponding feature maps using binary masks. During training, we also employ random modality dropout on ECG, EMG, and EOG with probability *p*_mod_ = 0.05, encouraging the model to retain performance in scenarios where some signals are absent or corrupted [26, 27]. Random modality dropout was not applied to EEG, as we assume during PSG acquisition at least some of the full montage EEG channels are present.

Fusion and temporal encoding: The outputs of the four modality-specific branches are concatenated along the feature dimension, yielding a representation of shape (*B,* 4 *× C, T*). A modality-level gating mechanism (sigmoid gating) then computes weights for each modality via a fully connected network operating on global averages. These weights are applied to rescale the per-modality contributions, allowing the network to emphasise the most informative signals on a per-sample basis. The weighted features are projected into a common embedding space and passed into a Temporal Context Encoder (TCE). The TCE and classification modules remain as in the original AttnSleep model.

## 4 Signal preprocessing

Preprocessing consisted of a global PSG cleaning stage applied to all recordings, followed by experiment-specific pipelines reflecting differing model and modality requirements. Subject inclusion therefore varied across experiments due to channel availability constraints.

### 4.1 Global PSG preprocessing

Due to multiple interventions on certain recording nights, a subset of the files contained discontinuities. Consequently, these recordings could not be analysed as continuous time series. Instead, they were truncated to the longest uninterrupted segment, provided it lasted at least five hours and began at the point sleep staging had commenced. For all other recordings, the data was originally truncated using the annotation files - from the point at which sleep staging commenced to the end of sleep staging. From this point, we describe the preprocessing steps for evaluation of the baseline models and the preprocessing steps for training and evaluation of AttnSleep and the multi-modal model.

### 4.2 Preprocessing: baseline models

For AttnSleep, we trained it on both datasets in the original paper (Sleep-EDF-78 and SHHS) to assess whether larger training data improved performance prior to testing on our cohort. The SHHS dataset (sampled at 125 Hz) was downsampled to 100 Hz to match the Sleep-EDF-78 dataset. Downsampling was performed using a polyphase FIR filter (SciPy *resamplepoly*, the nominal cut-off frequency corresponds to the new Nyquist frequency, with an effective transition band of 48-58 Hz. The same preprocessing steps were applied to the SW and MASS data, as in the original publications: signals were downsampled to 100 Hz and epochs labelled as unknown were removed. For AttnSleep, the EEG channel was selected according to availability, with preference given to Fpz–Cz, followed by C4–A1, as specified in [25]. For the RF model, EEG channel selection followed the preferential order specified in [19]: C4-A1, C3-A2 or C1-A1. As is common in clinical sleep staging, some recordings had missing modalities, either due to electrode detachment or exclusion during file export. To ensure a fair comparison between the RF and AttnSleep models, we restricted the analysis to the 89 subjects with complete EEG, EOG, and chin EMG recordings, which are required for sleep staging with the RF. One healthy control, one subject with RBD, and one subject with another sleep disorder were removed for incomplete recordings; a further subject with another sleep disorder was excluded at derivation as non-diagnostic. The final dataset is described in table 3.

**Table 3:** Dataset composition and sleep stage distribution across subject groups.

| Group | N Subj | N Epochs | Wake | N1 | N2 | N3 | REM |
| --- | --- | --- | --- | --- | --- | --- | --- |
| All | 142 | 142583 | 22.6% | 12.2% | 39.8% | 12.2% | 13.2% |
| SW Control | 18 | 17543 | 20.7% | 9.2% | 40.6% | 15.5% | 14.1% |
| MASS Control | 53 | 50989 | 23.4% | 13.9% | 43.5% | 6.7% | 12.5% |
| RBD | 31 | 32065 | 23.8% | 12.5% | 33.9% | 15.5% | 14.4% |
| Other Patient | 40 | 41986 | 21.4% | 11.0% | 39.6% | 15.1% | 12.9% |

### 4.3 Preprocessing: multimodal and single EEG comparison

To evaluate model performance on mixed-pathology clinical data, both the single-EEG and multimodal AttnSleep configurations were trained using the same combined dataset. SW subjects from Papworth were removed as they had no ECG trace, for fair comparison. The dataset comprised of 66 clinical SW subjects and 53 healthy controls from the MASS dataset. A summary of the sleep stage distribution across subjects is provided in table 4. All recordings were preprocessed using a consistent pipeline to ensure comparability across model configurations. To mitigate amplitude differences arising from heterogeneous recording equipment, electrode impedances, and amplifier gains across sites, all signals were z-score normalised on a per-subject basis prior to model input.

**Table 4:** Dataset composition and sleep stage distribution for the retrained cohort.

| Group | N Subj | N Epochs | Wake | N1 | N2 | N3 | REM |
| --- | --- | --- | --- | --- | --- | --- | --- |
| All | 119 | 117029 | 22.2% | 13.1% | 38.2% | 13.1% | 13.3% |
| SW Control | 13 | 11929 | 17.1% | 9.2% | 38.9% | 19.7% | 15.0% |
| MASS Control | 53 | 50989 | 23.4% | 13.9% | 43.5% | 6.7% | 12.5% |
| RBD | 22 | 22787 | 22.6% | 14.1% | 29.0% | 18.5% | 15.8% |
| Other Patient | 31 | 31324 | 21.9% | 12.6% | 36.0% | 17.2% | 12.3% |

As MASS recordings were already filtered and referenced, identical preprocessing steps were applied to the SW in-clinic data for consistency. The EEG signal was selected according to availability in the following preferential order: C3-CLE, C4-CLE, Fpz–CLE. EEG signals were referenced using a computed linked-ear reference. A 4th order Butterworth notch filter was applied at 50 Hz to remove powerline interference, and a 0.3 Hz high-pass 1st order Butterworth filter applied to attenuate baseline drift and remove phase distortion. Filtering was performed in a zero-phase manner to avoid phase distortion. All EEG signals were resampled to 100 Hz.

In regard to the data for the multimodal model, the same filtering and referencing procedures applied to the MASS dataset were also used for the SW data. Based on the overlap in available channels across recordings from both datasets, the following EEG channels were selected: C3, C4, O1, O2, Cz, F3, F4, F7, F8, Fz, P3, P4, Pz, T3, T4, T5, and T6. EEG signals were referenced using a computed linked-ear reference and filtered with a 1st order Butterworth high-pass filter at 0.3 Hz, along with a 50 Hz Butterworth band-stop notch filter to remove power-line interference. All EEG signals were downsampled to a target sampling frequency of 100 Hz.

For the ECG, a single channel in a D-I lead configuration was used, formed as a bipolar derivation between the left and right electrodes. The ECG signal was band-pass filtered using a 1st order Butterworth filter with cut-off frequencies of 0.1–30 Hz, combined with a 50 Hz notch filter, and downsampled to 200 Hz. The EOG signal was computed as the bipolar difference between the right and left outer canthi (ROC–LOC) and filtered using a 1st order Butterworth band-pass filter between 0.1 and 30 Hz, with an additional 50 Hz notch filter; the resulting signal was downsampled to 100 Hz. The chin EMG signal (submentalis) was also treated as a bipolar derivation and filtered using a first-order Butterworth band-pass filter between 10 and 100 Hz, along with a 50 Hz notch filter, before resampling to 200 Hz. All filters were applied in a zero-phase manner to avoid phase distortion.

## 5 Training and evaluation

For evaluation of the baseline models, we apply the RF and AttnSleep as is to our SW+MASS dataset (table 3). For the RF, we do not assess the performance on the MASS subjects, as the original model was trained on this cohort. We trained AttnSleep on the Sleep-EDF-78 + SHHS dataset, using the original hyperparameters, using the resulting model from a random fold as our baseline AttnSleep.

During training and evaluation of AttnSleep architecture on the SW+MASS dataset (table 4), we employed 10-fold cross-validation, with results subsequently aggregated. Subjects were stratified based on RBD vs. non-RBD, to ensure that the training and validation datasets were as consistent as possible across folds. For the multi-modal model the learning rate was reduced to 3e-4 to stabilise training, and all other hyperparameters were retained from the original set up. The models were trained for a maximum of 100 epochs for computational feasibility, with early stopping applied using a validation patience of 10 epochs. We compared the performance difference of the single-channel configuration the multi-modality configuration.

Beyond aggregate results, we analysed the interpretability of the multi-modal, focusing on the relative contribution of each sensing modality to different sleep stages. Modality weightings were extracted for each EEG channel, and each modality as a whole. Missing modalities did not contribute to the weighting analysis, as they were explicitly masked during model training and inference. EEG channel fusion employed a softmax attention mechanism, where a lightweight MLP scored each available channel and normalised weights across all present channels, ensuring the contributions summed to one. Channels absent from a given recording were masked prior to the softmax operation, causing their weight to be redistributed among the remaining channels. In contrast, modality-level fusion used a sigmoid gating mechanism, which assigned each modality an independent importance weight in the range of 0 to 1. Absent modalities were masked to negative infinity before the sigmoid activation, yielding a gate value of approximately zero, while the weights of the remaining modalities were unaffected. This design choice reflects the different roles of the two fusion stages: softmax channel attention enforces competition among EEG channels, encouraging the model to focus on the most informative derivations, while sigmoid modality gating allows complementary modalities (e.g., EEG and EOG) to contribute simultaneously at full capacity without suppressing one another. To account for potential bias arising from unequal channel availability across recordings, we also computed availability-corrected mean weights by averaging each channel’s (or modality’s) attention weight only over epochs in which it was present, excluding epochs where it was masked as absent.

To identify an optimal reduced-channel configuration, we leveraged the learned attention weights from the full multi-modal model as a data-driven channel and modality selection criterion. The top-ranked EEG channels and the non-EEG modality exhibiting the greatest discriminative contribution (particularly for underperforming clinical subgroups) were retained, and a new model was trained using only the selected inputs. This attention-guided pruning approach reduces computational cost and input requirements while preserving the most informative signal sources as determined by the model itself.

Model configurations were tested for significance versus the baseline single channel EEG AttnSleep model. To account for multiple pairwise comparisons across model configurations and cohorts, p-values were adjusted using the Holm-Bonferroni step-down procedure, controlling the family-wise error rate at *α*=0.05. Bootstrap confidence intervals (1000 resamples, subject-level) on the paired difference were computed in parallel. A comparison was considered statistically significant only when both the Holm-corrected p-value was below 0.05 and the bootstrap CI excluded zero.

## 6 Subgroup analysis

In all experiments, we evaluated performance within diagnostic subgroups and across sleep stages, to understand both population-specific limitations and stage-specific challenges. First, we compared individuals with RBD to healthy controls, reflecting the conventional case–control setting commonly used in prior work to characterise disease-specific effects. Second, to better approximate real-world clinical deployment, we compared individuals with RBD to a broader non-RBD group comprising both healthy controls and patients with other sleep disorders. This latter comparison reflects the heterogeneous population typically referred for in-clinic polysomnography, where the practical diagnostic task is to distinguish RBD from a wide range of alternative conditions rather than from healthy individuals alone.

## 7 Evaluation metrics

We evaluate the model using three evaluation metrics: accuracy, Cohen’s Kappa, and the mean F1 score. Cohen’s *κ* adjusts for agreement expected by chance, making it more comparable to human inter-scorer agreement. Clinically, a higher *κ* indicates the model is approaching the consistency expected between trained scorers, with a value of 1 indicating perfect agreement. Here it is the agreement between the expert rater and the model:

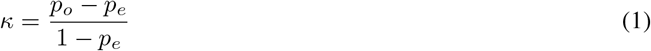

where: *p_o_*= observed agreement, *p_e_*= expected agreement by chance.

The F1 score is the harmonic mean of precision and recall, particularly useful when dealing with class imbalance, and is defined as:

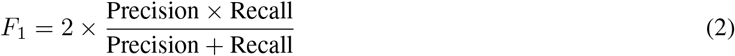

where: 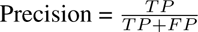, 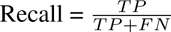. (TP: True positives, FP: False positives, FN: False negatives). The mean F1 when dealing with multiple classes is given as:

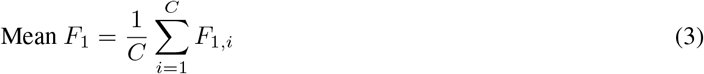

where: 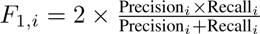, and *C* is the total number of classes.

Accuracy is given as:

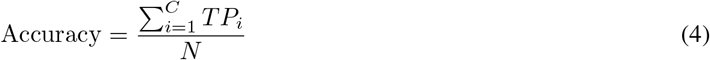

where: *TP_i_* is the number of correctly classified samples for class *i*, *C* is the total number of classes, and *N* is the total number of classes.

## 8 Results

### 8.1 Baseline models

The performance of AttnSleep when trained on each control dataset individually and the combined Sleep-EDF-78 + SHHS is summarized in table 5, alongside results from individual datasets for comparison. When trained on the combined dataset (Sleep-EDF-78+SHHS), the model achieved a Cohen’s *κ* of 0.75. The performance of the model stratified by sleep stage and age bracket in shown in fig. 2. The distribution of subjects acrossage is given in the supplementary material (fig. 5). To evaluate generalisation, a model from a randomly selected fold was applied to the SW+MASS dataset (table 3). The RF model was evaluated on the same dataset for consistency.

**Figure 2:**
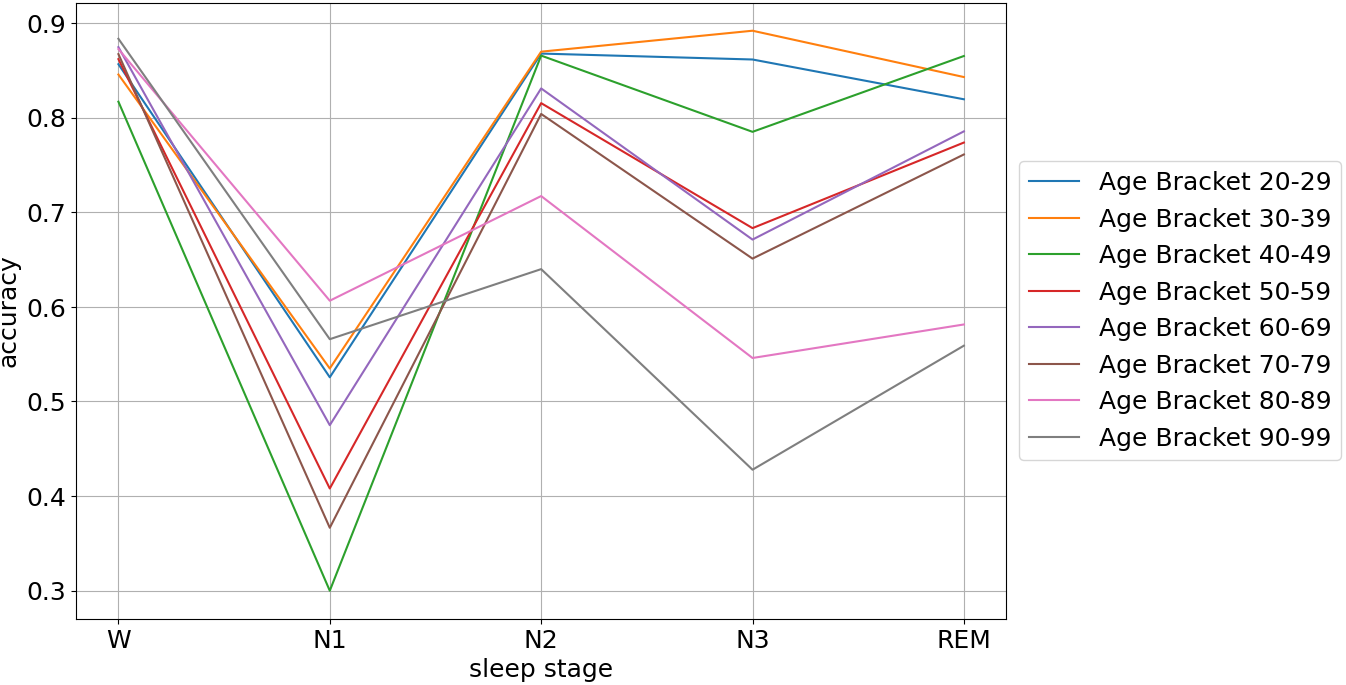
Accuracy of the AttnSleep model across sleep stages within age brackets within the combined SHHS and SleepEDF-78 dataset.

**Table 5:** AttnSleep: Performance metrics across different datasets.

|  | Accuracy | Mean F1 | Kappa |
| --- | --- | --- | --- |
| Sleep-EDF-20 | 84.2 | 78.6 | 0.78 |
| Sleep-EDF-78 | 80.7 | 75.7 | 0.73 |
| SHHS | 82.6 | 74.3 | 0.76 |
| Sleep-EDF-78 + SHHS | 81.1 | 75.5 | 0.75 |

The performance of the models in the SW+MASS dataset is summarised in table 6. The performance metrics are given for the SW cohort overall and by subgroup; MASS controls were evaluated for AttnSleep only, as the RF was not applied to that dataset. The RF achieved a *κ* of 0.56 and AttnSleep 0.49 overall, and in the RBD group the RF and Attnsleep achieved a *κ* of 0.54 and 0.45, respectively. Table 7 reports the *κ* metric for each sleep stage across the diagnosis subgroups using the RF and AttnSleep model. For REM in RBD subjects, the RF model achieved a *κ* of 0.51, whereas AttnSleep obtained 0.18.

**Table 6:** Results of each of the models on the SW dataset. Accuracy is 5-class accuracy for both models. Bold indicates the best value within each metric.

| Cohort | Accuracy |  | Mean F1 |  | Kappa |  |
| --- | --- | --- | --- | --- | --- | --- |
|  | RF | AttnSleep | RF | AttnSleep | RF | AttnSleep |
| SW overall | **67.4** | 62.1 | **60.5** | 56.6 | **0.56** | 0.49 |
| Controls SW | **67.3** | 63.0 | **60.2** | 57.1 | **0.55** | 0.50 |
| Controls MASS | - | 75.4 | - | 70.4 | - | 0.66 |
| non-RBD | **68.4** | 64.0 | **61.3** | 58.3 | **0.57** | 0.51 |
| RBD | **65.3** | 58.4 | **59.0** | 53.4 | **0.54** | 0.45 |

**Table 7:** Cohen’s *κ* per sleep stage across diagnosis groups for RF and AttnSleep.

| Sleep Stage | Random Forest |  |  | AttnSleep |  |  |
| --- | --- | --- | --- | --- | --- | --- |
|  | SW HC | Non-RBD | RBD | SW HC | Non-RBD | RBD |
| W | 0.71 | 0.67 | 0.64 | 0.64 | 0.63 | 0.58 |
| N1 | 0.22 | 0.20 | 0.17 | 0.16 | 0.22 | 0.22 |
| N2 | 0.54 | 0.57 | 0.55 | 0.56 | 0.57 | 0.53 |
| N3 | 0.55 | 0.62 | 0.64 | 0.64 | 0.65 | 0.63 |
| REM | 0.56 | 0.59 | 0.51 | 0.38 | 0.37 | 0.18 |

### 8.2 Multimodal model and single EEG comparison

This section evaluates whether incorporating multiple PSG modalities provides a measurable performance advantage over a single-channel EEG configuration. We compare the multi-modal AttnSleep model against its original single-EEG counterpart using identical subject splits, enabling an assessment of the contribution of additional modalities. Performance is reported as the mean *±* standard deviation across cross-validation folds, as summarised in table 8. The multi-modal model improved performance in the cohort overall (*κ* 0.61 versus 0.63), in healthy controls (*κ* 0.68 versus 0.70) and in non-RBD subjects (*κ* 0.63 versus 0.66). In RBD subjects, however, performance was essentially unchanged (*κ* 0.46 for both configurations, fold-mean values), indicating that the benefit of the additional modalities did not extend to the RBD subgroup.

**Table 8:** Performance comparison between the baseline 1-EEG model and the Multimodal model (mean *±* SD).

| Group | Model | Accuracy | Mean F1 | Cohen’s $\kappa$ |
| --- | --- | --- | --- | --- |
| All | 1-EEG | 70.68 ± 3.53 | 66.57 ± 3.39 | 0.61 ± 0.05 |
|  | Multimodal | **72.09 ± 3.35** | **68.94 ± 3.75** | **0.63 ± 0.05** |
| HC | 1-EEG | 76.11 ± 3.50 | 71.38 ± 2.97 | 0.68 ± 0.04 |
|  | Multimodal | **78.18 ± 2.47** | **74.32 ± 2.27** | **0.70 ± 0.03** |
| Non-RBD | 1-EEG | 72.72 ± 3.53 | 68.10 ± 3.23 | 0.63 ± 0.05 |
|  | Multimodal | **74.30 ± 3.20** | **70.77 ± 3.63** | **0.66 ± 0.04** |
| RBD | 1-EEG | **58.64 ± 13.44** | 52.83 ± 16.31 | **0.46 ± 0.19** |
| | Multimodal | 58.45 ± 15.53 | **54.15 ± 17.47** | **0.46 ± 0.21** |

In addition to overall performance metrics, we compared the stage-specific performance. Table 9 shows Cohen’s *κ* for the single-EEG and multi-modal models in each subgroup. In the overall cohort and in healthy controls, the multi-modal model achieved higher *κ* in every stage, with the largest gains in N1 (*κ* 0.32 versus 0.36 overall; 0.40 versus 0.45 in healthy controls) and N3 (*κ* 0.64 versus 0.68 overall; 0.65 versus 0.71 in healthy controls). The same pattern held in non-RBD subjects for all stages except wake, where the two models were equivalent (*κ* 0.80). In RBD subjects the pattern differed: the gain was concentrated in wake (*κ* 0.63 versus 0.70), while N2 (*κ* 0.49 versus 0.45) and N3 (*κ* 0.68 versus 0.67) were not improved, and REM rose only modestly (*κ* 0.53 versus 0.55).

**Table 9:** Stage-level Cohen’s *κ* comparison between 1-EEG and Multimodal models across subject subgroups.

| Stage | All |  | HC |  | Non-RBD |  | RBD |  |
| --- | --- | --- | --- | --- | --- | --- | --- | --- |
|  | 1-EEG | Multi | 1-EEG | Multi | 1-EEG | Multi | 1-EEG | Multi |
| Wake | 0.76 | **0.78** | 0.86 | **0.87** | **0.80** | **0.80** | 0.63 | **0.70** |
| N1 | 0.32 | **0.36** | 0.40 | **0.45** | 0.34 | **0.39** | 0.23 | **0.24** |
| N2 | 0.60 | **0.61** | 0.67 | **0.70** | 0.62 | **0.65** | **0.49** | 0.45 |
| N3 | 0.64 | **0.68** | 0.65 | **0.71** | 0.63 | **0.68** | **0.68** | 0.67 |
| REM | 0.63 | **0.66** | 0.68 | **0.72** | 0.66 | **0.68** | 0.53 | **0.55** |

### 8.3 Modality contribution

Here we report the importance of each of the channels included in the multi-modality model prior to channel fusion. These were averaged across all folds. Table 10 a) shows the weighting of each of the group modalities overall and across each sleep stage and b) shows the contribution of different modalities across RBD subjects compared to subjects without RBD. Across all subjects, EEG received the highest weighting (0.312 overall).EOG contributed most strongly during REM and N1 epochs (0.223 and 0.222 respectively) and least during N3 (0.172), consistent with the ocular dynamics of these stages. ECG was most influential during N3 (0.301), where it slightly exceeded the EEG weighting (0.299). In RBD subjects, EOG showed a higher weighting relative to subjects without RBD (0.273 versus 0.177, Δ = +0.096), while EEG weighting fell (Δ = *−*0.056) and EMG weighting decreased slightly (Δ = *−*0.011).

**Table 10:** Modality importance analysis. (a) Modality weights across sleep stages for all subjects. (b) Comparison between RBD and subjects without RBD (Δ = RBD *−* without RBD).

| Stage | EEG | ECG | EMG | EOG |
| --- | --- | --- | --- | --- |
| Overall | 0.312 | 0.244 | 0.173 | 0.196 |
| Wake | 0.377 | 0.242 | 0.196 | 0.192 |
| N1 | 0.301 | 0.216 | 0.159 | 0.222 |
| N2 | 0.290 | 0.246 | 0.156 | 0.188 |
| N3 | 0.299 | 0.301 | 0.192 | 0.172 |
| REM | 0.287 | 0.210 | 0.177 | 0.223 |

(b) RBD vs Without RBD
|  | EEG | ECG | EMG | EOG |
| --- | --- | --- | --- | --- |
| RBD | 0.267 | 0.249 | 0.164 | 0.273 |
| Without RBD | 0.323 | 0.243 | 0.175 | 0.177 |
| $\Delta$ (RBD – Without RBD) | −0.056 | 0.006 | −0.011 | **+0.096** |

Figure 3 shows the weighting of each of the EEG channels in the EEG MRCNN branch, across each sleep stage. Central channels (Cz, C4, C3) had the highest weighting across N1, N2, N3, and REM, whereas channels exhibited more uniform weighting in wake. The parietal P3 and P4 consistently showed low weighting across all stages.

**Figure 3:**
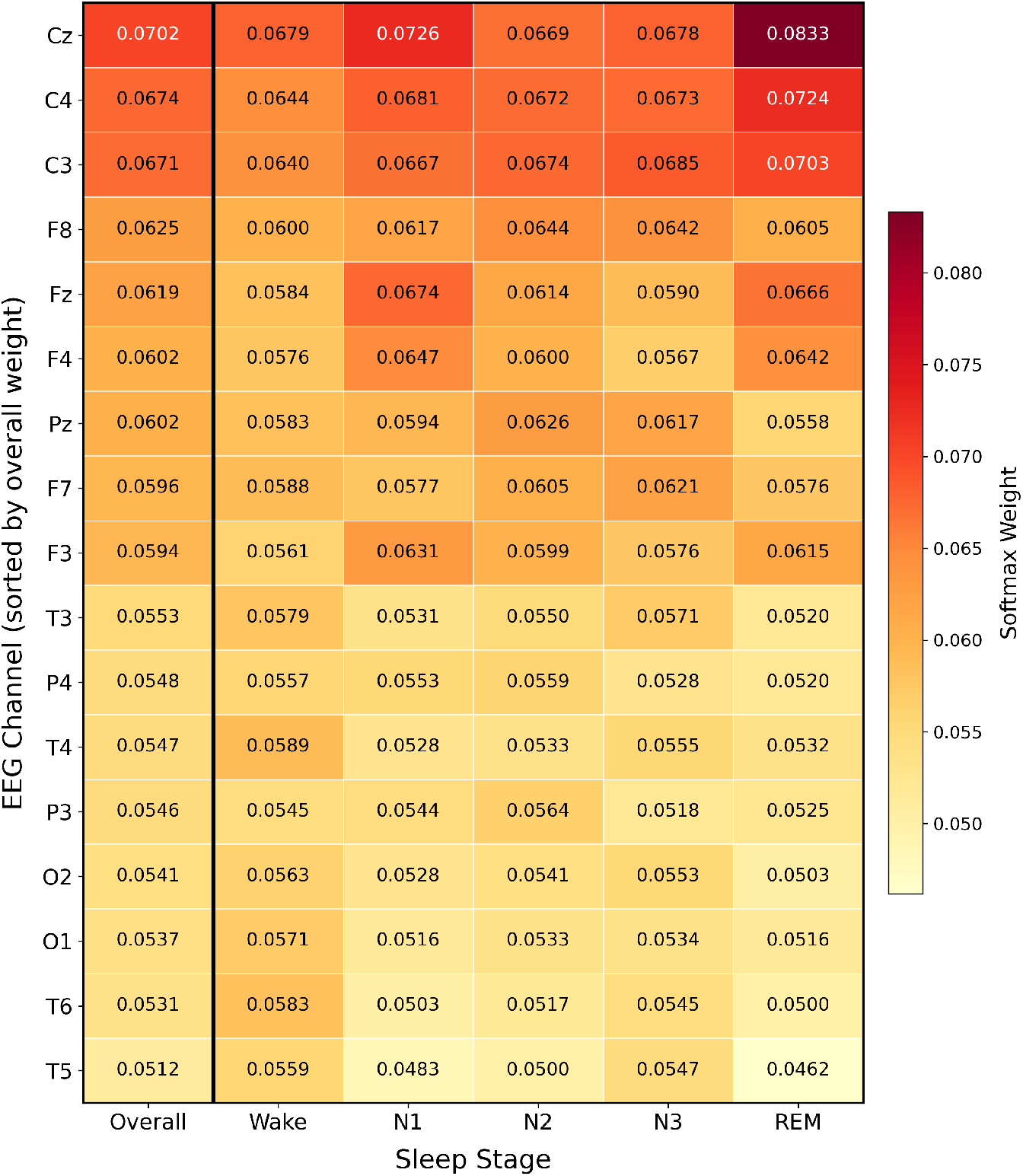
Learned average weighting for each of the EEG channels included in the multi-modality model. Adjusted for bias in relation to missing channels.

### 8.4 Targeted multi-modal expansion

To investigate whether a reduced-channel model could match or exceed the full multi-modal configuration, we examined the learned attention weights to guide architecture selection. EEG consistently received the highest modality weight across all sleep stages (mean = 0.31), confirming it as the dominant modality for sleep staging. We therefore first evaluated a 4-channel EEG model using the channels with the highest raw attention weights: C3, C4, Cz, and F8. This model achieved comparable performance to the full 17-channel multi-modal model while requiring substantially fewer input channels.

To determine whether a complementary non-EEG modality could further improve performance, we compared the modality attention weights between RBD and non-RBD subjects. EOG showed the largest weight increase in RBD subjects (0.273 vs 0.177 in non-RBD, Δ = +0.096), consistent with the preservation of rapid eye movements during REM sleep in RBD despite disrupted EEG and EMG patterns. In contrast, ECG was unchanged (Δ = +0.006) and EMG decreased (Δ = *−*0.011) and contributed to overall performance degradation in the full multi-modal model, particularly for RBD subjects where EMG atonia patterns are fundamentally altered. We therefore constructed a final model combining the 4 selected EEG channels with EOG. The results for all models are compared in table 11. The 4 EEG + EOG model matched the full multi-modal model in overall agreement (*κ* = 0.63 for both) despite using a substantially reduced channel set, and achieved the highest *κ* of any configuration in RBD subjects (*κ* = 0.49).

**Table 11:** Performance comparison across all evaluated model configurations (mean *±* SD).

| Group | Model | Accuracy | Mean F1 | Cohen’s $\kappa$ |
| --- | --- | --- | --- | --- |
| All | 1-EEG | 70.68 ± 3.53 | 66.57 ± 3.39 | 0.61 ± 0.05 |
| | Multimodal | 72.09 ± 3.35 | 68.94 ± 3.75 | 0.63 ± 0.05 |
| | 4-EEG | 71.28 ± 3.66 | 67.69 ± 3.61 | 0.62 ± 0.05 |
|  | 4-EEG + EOG | **72.13 ± 3.47** | **69.03 ± 3.68** | **0.63 ± 0.05** |
| Healthy | 1-EEG | 76.11 ± 3.50 | 71.38 ± 2.97 | 0.68 ± 0.04 |
|  | Multimodal | **78.18 ± 2.47** | **74.32 ± 2.27** | **0.70 ± 0.03** |
| | 4-EEG | 76.72 ± 4.30 | 72.55 ± 3.99 | 0.69 ± 0.05 |
| | 4-EEG + EOG | 77.50 ± 3.30 | 73.47 ± 3.26 | 0.70 ± 0.04 |
| Non-RBD | 1-EEG | 72.72 ± 3.53 | 68.10 ± 3.23 | 0.63 ± 0.05 |
|  | Multimodal | **74.30 ± 3.20** | **70.77 ± 3.63** | **0.66 ± 0.04** |
| | 4-EEG | 73.43 ± 4.04 | 69.48 ± 4.05 | 0.65 ± 0.05 |
| | 4-EEG + EOG | 74.05 ± 3.59 | 70.53 ± 3.71 | 0.65 ± 0.05 |
| RBD | 1-EEG | 58.64 ± 13.44 | 52.83 ± 16.31 | 0.46 ± 0.19 |
| | Multimodal | 58.45 ± 15.53 | 54.15 ± 17.47 | 0.46 ± 0.21 |
| | 4-EEG | 59.26 ± 13.60 | 53.01 ± 16.38 | 0.46 ± 0.20 |
|  | 4-EEG + EOG | **61.39 ± 12.76** | **55.59 ± 16.68** | **0.49 ± 0.19** |

The stage wise performance in shown in table 12 for the single EEG, multi-modal model, and the 4 EEG + EOG configuration. The latter achieved the best REM classification with a mean F1 of 73.5, compared to 68.3 using the single EEG model. In RBD subjects the 4 EEG + EOG configuration also had the highest REM performance (60.6% recall), outperforming both the single-channel EEG and full multi-modal configurations, as shown in the confusion matrices in **??**.

**Table 12:** Stage-wise performance comparison across model configurations (mean F1).

| Stage | Single EEG | Multi-modal | 4 EEG + EOG |
| --- | --- | --- | --- |
| Wake | 81.9 | **83.0** | 82.1 |
| N1 | 39.4 | 44.3 | **44.8** |
| N2 | 74.9 | **75.7** | 75.5 |
| N3 | 68.9 | **71.9** | 69.9 |
| REM | 68.3 | 70.8 | **73.5** |
| All | 66.7 | **69.1** | **69.1** |

We also report the pooled performance metrics in the SW controls versus the controls from the MASS dataset in the supplementary material table 22, to examine the performance discrepancy between cohorts.

### 8.5 Statistical comparison across configurations

To assess whether observed differences between model configurations were statistically meaningful, we computed 95% bootstrap confidence intervals (2000 subject-level resamples) on per-subject Cohen’s *κ* and macro-F1, and performed Holm-corrected paired Wilcoxon signed-rank tests on pairwise model comparisons. Results are summarised in tables 13 and 14.

**Table 13:** Cohen’s *κ* and macro-F1 across model configurations, reported as pooled mean across folds with 95% bootstrap confidence intervals (2000 resamples, subject-level). Best value per row in bold.

| Metric | Cohort | Single-EEG | Multi-modal | 4-EEG + EOG |
| --- | --- | --- | --- | --- |
| Cohen’s $\kappa$ | Overall (n=119) | 0.61 [0.58, 0.64] | **0.63** [0.60, 0.66] | **0.63** [0.60, 0.66] |
|  | RBD (n=22) | 0.53 [0.45, 0.60] | 0.54 [0.46, 0.61] | **0.56** [0.48, 0.62] |
|  | Non-RBD (n=97) | 0.63 [0.60, 0.66] | **0.65** [0.62, 0.69] | **0.65** [0.62, 0.68] |
| Macro-F1 | Overall (n=119) | 0.67 [0.64, 0.69] | **0.69** [0.66, 0.72] | **0.69** [0.67, 0.71] |
|  | RBD (n=22) | 0.60 [0.54, 0.65] | 0.62 [0.55, 0.67] | **0.63** [0.57, 0.68] |
|  | Non-RBD (n=97) | 0.68 [0.65, 0.70] | **0.71** [0.68, 0.74] | 0.70 [0.68, 0.73] |

**Table 14:** Pairwise statistical comparisons between model configurations. Holm-corrected *p*-values on per-subject scores.

| Metric | Comparison | Overall | RBD | Non-RBD |
| --- | --- | --- | --- | --- |
| Cohen’s $\kappa$ | Single-EEG vs Multi-modal | $8 \times 10^{-3}$ | 0.78 | $5 \times 10^{-3}$ |
| | Single-EEG vs 4-EEG + EOG | $2 \times 10^{-4}$ | 0.18 | $1 \times 10^{-3}$ |
| Macro-F1 | Single-EEG vs Multi-modal | $7 \times 10^{-4}$ | 0.63 | $5 \times 10^{-4}$ |
| | Single-EEG vs 4-EEG + EOG | $1 \times 10^{-8}$ | 0.07 | $1 \times 10^{-7}$ |

For Cohen’s *κ*, the multi-modal configuration significantly outperformed the single-EEG baseline in the overall cohort (Holm *p* = 8 *×* 10*^−^*^3^) and in non-RBD subjects (Holm *p* = 5 *×* 10*^−^*^3^), but not in RBD subjects (Holm *p* = 0.78). The 4-EEG + EOG configuration likewise outperformed the baseline overall (Holm *p* = 2 *×* 10*^−^*^4^) and in non-RBD subjects (Holm *p* = 1 *×* 10*^−^*^3^), with bootstrap confidence intervals on the paired difference excluding zero in both cases, but the corresponding comparison in RBD subjects was not significant (Holm *p* = 0.18). For macro-F1 the same pattern held: both configurations significantly outperformed the baseline overall and in non-RBD subjects (all *p ≤* 7 *×* 10*^−^*^4^), while neither reached significance in RBD subjects (*p* = 0.63 for multi-modal; *p* = 0.07 for 4-EEG + EOG). No comparison within the RBD subgroup reached statistical significance, which we interpret in light of the limited RBD sample size.

**Figure 4:**
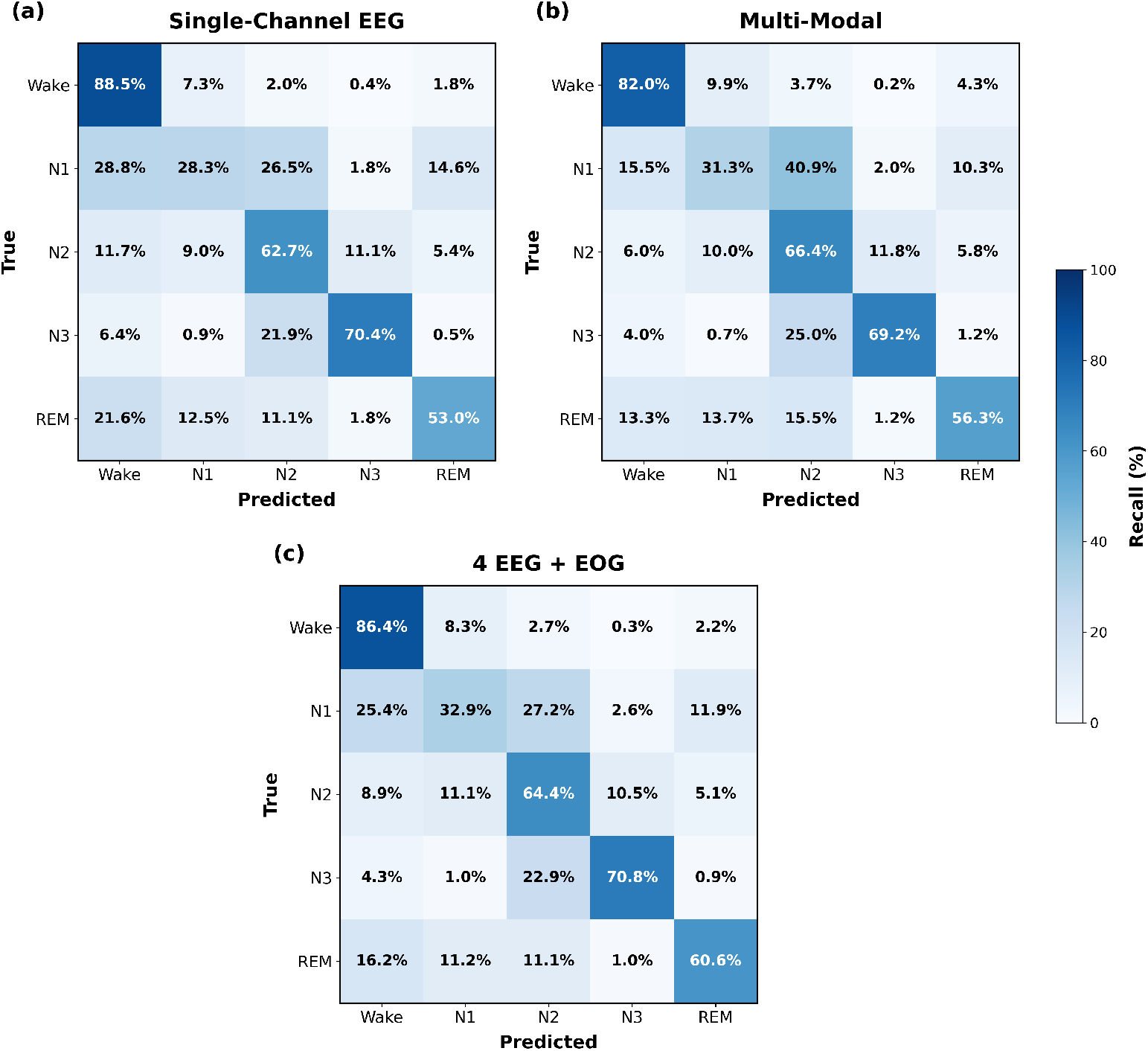
Confusion matrices for sleep stage classification in RBD subjects (n = 22) across three model configurations. Values represent per-class recall (%).

We then examined REM classification performance specifically within the RBD subgroup (tables 15 and 16). REM *κ* in RBD subjects was 0.53 [0.42, 0.61] for the single-EEG model, 0.55 [0.42, 0.66] for the multi-modal model, and 0.60 [0.49, 0.69] for the 4-EEG + EOG configuration. Only the 4-EEG + EOG configuration significantly outperformed the single-EEG baseline (Δ*κ* = *−*0.07, Holm *p* = 0.03, 95% CI [*−*0.11, *−*0.03]); the multi-modal configuration did not (Δ*κ* = *−*0.02, Holm *p* = 0.26, CI [*−*0.07, +0.03]). The same pattern was observed for REM F1, with the 4-EEG + EOG model achieving 0.66 [0.55, 0.74] compared with 0.59 [0.49, 0.67] for single-EEG (ΔF1 = *−*0.06, Holm *p* = 0.05), while the multi-modal comparison remained non-significant (*p* = 0.49). REM recall in RBD subjects rose from 53.0% with single-EEG to 56.3% with multi-modal and 60.6% with 4-EEG + EOG; REM precision rose from 67.0% to 67.5% and 71.4% respectively.

**Table 15:** REM sleep classification performance in RBD subjects (*n* = 22), pooled across folds. Values shown as mean with 95% bootstrap confidence intervals.

| Metric | Single-EEG | Multi-modal | 4-EEG + EOG |
| --- | --- | --- | --- |
| REM recall (%) | 53.0 [40.5, 63.6] | 56.3 [42.7, 68.8] | **60.6** [48.7, 70.8] |
| REM precision (%) | 67.0 [56.3, 75.2] | 67.5 [55.4, 78.0] | **71.4** [62.1, 78.2] |
| REM F1 | 0.59 [0.49, 0.67] | 0.61 [0.49, 0.71] | **0.66** [0.55, 0.74] |
| REM $\kappa$ (1-vs-rest) | 0.53 [0.42, 0.61] | 0.55 [0.42, 0.66] | **0.60** [0.49, 0.69] |

**Table 16:** Pairwise statistical comparisons of REM classification performance in RBD subjects (*n* = 22). Holm-corrected *p*-values and 95% bootstrap confidence intervals on the paired difference (configuration *−* single-EEG baseline).

| Metric | Comparison | $\Delta$ | Holm $p$ | 95% CI |
| --- | --- | --- | --- | --- |
| REM $\kappa$ | Single-EEG vs Multi-modal | -0.02 | 0.26 | [-0.07, +0.03] |
|  | Single-EEG vs 4-EEG + EOG | -0.07 | 0.03 | [-0.11, -0.03] |
| REM F1 | Single-EEG vs Multi-modal | -0.02 | 0.49 | [-0.06, +0.03] |
|  | Single-EEG vs 4-EEG + EOG | -0.06 | 0.05 | [-0.10, -0.03] |

## 9 Discussion

In this work, we evaluated the performance and generalisability of two representative approaches to automatic sleep staging within a mixed cohort (SW+MASS), including patients with RBD: a classical RF model and a state-of-the-art deep learning model, AttnSleep. When applied out of the box to our multi-site in-clinic PSG dataset, the RF model had a similar performance to the deep learning approach across the SW+MASS cohort as a whole, but outperformed AttnSleep within the RBD subgroup specifically. However, agreement with manual scoring, as measured by Cohen’s kappa, remained “moderate” according to Cohen’s *κ* benchmarks (0.41-0.60) [28]. In RBD subjects, we saw the lowest agreement overall in both models, with a Cohen’s *κ* of 0.43 in REM when applying the pre-trained RF. A meta-analysis of inter-rater variability in sleep staging found that manual rater agreement across studies was 0.76 overall, but varied between sleep stages, with Cohen’s *κ* values of 0.70, 0.24, 0.57, 0.57, and 0.69 for wake, N1, N2, N3, and REM, respectively [29]. Therefore, performance in the RBD subgroup remained below what can be considered clinically applicable thresholds. This underscores the inherent difficulty of generalising models trained on healthy or homogeneous cohorts, while also demonstrating the value of disease-specific adaptation.

The RF model was originally trained on a combination of controls and RBD patients, whereas AttnSleep was trained solely on healthy controls and relies on a single EEG channel as input. These results therefore not only reinforce prior findings that RBD presents a unique challenge for automated staging due to its atypical REM physiology, but also underscore the need for more diverse and representative training data to achieve generalisation. Before moving decisively toward minimal-sensor setups, it is essential to establish which combinations of modalities (e.g., EEG, EOG, EMG) contribute most to robust model performance, especially in heterogeneous clinical populations such as RBD. The influence of dataset composition is further evident when evaluating AttnSleep in control subjects. As shown in fig. 2, model performance declines progressively with age, particularly for REM and N3 stages, reflecting the model’s difficulty in generalising to more fragmented and abnormal sleep architectures commonly observed in older adults [30]. This trend is closely linked to the age distribution of the training set fig. 5), which is heavily skewed toward middle-aged subjects, with 50–59 year-olds comprising the largest cohort (n=100) and the 40–59 range accounting for nearly half of all subjects. In contrast, the oldest brackets are substantially under-represented (n=21 for 80–89 and n=12 for 90–99), and this imbalance is mirrored in the accuracy curves: the 80–89 and 90–99 groups show markedly lower performance on N3 (0.55 and 0.43, respectively) and REM (0.58 and 0.56), whereas the 30–49 brackets, which dominate training, achieve the highest accuracies across most stages. The 20–29 bracket performs comparably to the dominant middle-aged cohorts despite its small size (n=23), indicating that under-representation alone does not determine accuracy. Rather, younger subjects benefit from more regular, well-defined sleep architecture that generalises readily from limited samples, whereas older subjects are penalised by the combination of sparse training data and greater physiological variability. Previous studies have shown that sleep staging model performance varies across datasets, data quality, and patient population - all of which tend to be the case in clinical sleep staging [23, 31, 32]. Classical feature-based approaches such as RFs also typically plateau at a lower ceiling compared to deep learning architectures when trained on sufficiently large datasets, due to their limited representational capacity [33]. Although we were unable to assess the impact of training the AttnSleep model on a larger clinical dataset, we extended its architecture to incorporate additional modalities in order to capture more complex electrophysiological interactions. Specifically, we added multi-resolution convolutional (MRCNN) modules for feature extraction across all available EEG channels, as well as chin EMG, EOG, and a single ECG lead. This multimodal extension improved overall performance in the cohort as a whole, in healthy controls and in non-RBD subjects, but not in RBD subjects, in whom overall agreement was unchanged. As illustrated in table 9, in the former groups the multi-modal model enhanced performance in every sleep stage, with the most pronounced improvements in N1 and N3 (*κ* 0.32 versus 0.36 and 0.64 versus 0.68 respectively in the overall cohort), followed by REM (0.63 versus 0.66). In RBD subjects the pattern differed: gains were confined to wake and, more modestly, REM, while N2 and N3 showed slight declines. The stages showing the largest gains are those in which the inclusion of complementary modalities is physiologically justified, namely ECG in N3 and EOG in REM and N1.

A central question of this work was whether incorporating additional PSG modalities beyond a single EEG channel improves sleep staging in clinical populations, and in particular in RBD. At the aggregate cohort level, the full multi-modal model significantly outperformed the single-EEG baseline (Cohen’s *κ* 0.63 versus 0.61 overall (pooled over folds); Holm-corrected *p* = 8 *×* 10*^−^*^3^), but this advantage did not extend to the RBD subgroup, where overall agreement was unchanged (*κ* 0.54 versus 0.53; Holm *p* = 0.78).

Taken in isolation, this might suggest that the additional modalities offer little practical benefit. However, stage-stratified analysis shows that this apparent absence of benefit in RBD masks a substantial change in error structure. The multi-modal model approximately halved the rate at which sleep epochs were misclassified as wake in RBD subjects: N1*→*wake fell from 28.8% to 15.5%, N2*→*wake from 11.7% to 6.0%, REM*→*wake from 21.6% to 13.3%, and N3*→*wake from 6.4% to 4.0%. Consequently wake *κ* rose from 0.63 to 0.70 despite a fall in wake recall (88.5% to 82.0%), reflecting a gain in precision rather than sensitivity, and both N2 and REM recall improved (62.7% to 66.4% and 53.0% to 56.3% respectively). These gains were, however, offset by a corresponding increase in within-sleep confusion, principally N1*→*N2 (26.5% to 40.9%) and N3*→*N2 (21.9% to 25.0%). In effect, the additional modalities exchange a systematic tendency to over-call wake for a tendency to over-call N2, leaving overall agreement almost unchanged. This distinction matters clinically. Confidently mislabelling sleep as wake is the error mode that most distorts summary measures derived from staging, such as total sleep time and sleep efficiency.

These findings prompted a closer examination of modality-specific contributions to understand which signals were most informative for staging. The modality weights extracted from the fusion layer (table 10) indicate that EEG features carry the greatest overall importance across all stages. We observe a rise in EOG weighting during N1 and REM, and an increase in ECG contribution during N3 and wake. Although EEG dominates partly because multiple channels contribute to its feature representation, this is also consistent with the well-established central role of EEG in sleep staging through identification of characteristic changes in frequency bands such as alpha attenuation during sleep onset, spindle and K-complex activity in N2, and delta power in N3 [34, 35]. In N3, the observed increase in ECG weighting likely reflects the physiological drop in heart rate and heart rate variability that accompanies slow-wave sleep (SWS) [36, 37]. EOG remains a tertiary contributor across stages, with the lowest contribution in N3, which aligns with expected ocular dynamics across non-slow wave stages: rapid eye movements in REM, and transitional blinks or slow rolling movements in lighter sleep stages and in correspondence with arousals [35]. By contrast, the weighting for chin EMG remains consistently low. While expert scorers often rely on EMG tone reduction to identify REM, our RBD-inclusive cohort typically lacks the physiological REM atonia. Thus, the low reliance on EMG in the model is desirable, suggesting that REM staging was not driven by atonia cues that are unreliable or absent in RBD patients.

This finding is consistent with the modality-attention analysis. The model assigned increased EOG weighting during REM (*w* = 0.42 overall) and in RBD subjects specifically (*w* = 0.48 vs 0.40 in non-RBD, Δ = +0.08; Table 10). In RBD, where chin EMG no longer provides a reliable REM signature due to RSWA, the model appears to compensate by leaning more heavily on EOG, a modality that captures rapid eye movements, which are preserved in RBD despite the disrupted EMG and partially altered EEG patterns. The attention weights thus identify both why the multi-modal model helps in REM and why the gain is amplified in RBD, supporting the interpretation that the additional signals carry disease-relevant information rather than simply increasing model capacity.

The EEG channel-specific weighting also aligns with the neurophysiology of sleep staging. As shown in fig. 3, the model assigns the greatest importance to central electrodes, which dominate across most sleep stages. This is physiologically appropriate, as central derivations are traditionally used to capture fast sleep spindles and K-complexes, both of which are characteristic markers of stage N2 sleep [38, 39]. Parietal (Pz) and frontal electrodes exhibit moderate weighting, consistent with their roles in capturing distinct sleep-related rhythms: frontal channels tend to reflect delta activity during stage N3 and theta activity during REM sleep, while the posterior parietal sites help track alpha attenuation from wake to N1 and can also reflect spindles during N2 [40, 41, 38]. In contrast, the temporal electrodes contribute minimally, which aligns with manual scoring where temporal leads are less frequently used due to their lower discriminative value [35].

Some aspects, however, deviate slightly from classical manual scoring conventions. For example, while REM sleep EEG is typically dominated by theta activity distributed across frontal and central regions, the model shows a stronger emphasis on the central midline (Cz) alone [35]. Finally, although frontal slow waves are physiologically a prominent feature of stage N3, the model’s weights indicate stronger reliance on central rather than frontal sites, which typically capture slow spindles [38]. One plausible explanation is the relative robustness of the central midline electrode. Compared with frontal sites, Cz is typically less affected by eye and facial muscle artefacts, which can improve its effective signal-to-noise ratio [42]. Moreover, Cz reflects activity from a spatially broad cortical region due to its placement, potentially increasing utility.

By selecting the four EEG channels with the highest weights and using only these as input to a reduced model, we observed improved *κ* agreement relative to the single-EEG baseline in all groups (table 11). Incorporating the non-EEG modality associated with the greatest gain in RBD subjects (EOG), the 4-EEG + EOG model achieved the highest *κ* in the overall cohort (0.63, matching the full multi-modal model) and in RBD subjects (0.49), while the full multi-modal model achieved the highest agreement in healthy controls and non-RBD subjects. At the stage level, the 4-EEG + EOG model achieved the highest performance in REM, whereas the multi-modal model was best in wake, N2 and N3 (table 12), the latter likely reflecting the contribution of ECG. Notably, REM classification in RBD subjects improved, with the strongest performance observed for the 4-EEG + EOG model, followed by the multi-modal model (**??**). The 4-EEG + EOG configuration significantly outperformed the single-EEG baseline overall (Holm *p* = 2 *×* 10*^−^*^4^) and on REM *κ* in RBD (Holm *p* = 0.03), with the bootstrap CI on the paired difference excluding zero in both cases. The benefit of the targeted EOG addition is therefore concentrated where it is clinically most consequential, namely REM classification in the population in which standard atonia-based REM markers are unreliable. Notably, this is specific to EOG augmentation rather than to multimodality in general: the gains from the full multi-modal model reached significance in the overall and non-RBD cohorts but not within the RBD subgroup, where its improvement was confined to wake classification.

This reinforces the modality-attention findings: the contributions of ECG and EMG, while small on aggregate, may introduce noise that partially offsets the benefit of additional information; particularly in RBD, where EMG patterns deviate from training assumptions. Informed, data-driven modality selection therefore appears to outperform both minimal-sensor and full-multimodal setups.

Channel selection was based on raw (uncorrected) attention weights rather than availability-corrected weights. While corrected weights better reflect the model’s learned preference for each channel when present, raw weights additionally account for channel availability across the cohort, implicitly penalising channels that are absent in a subset of recordings. For a reduced-channel model intended to generalise across heterogeneous recording setups, this is a desirable property: selecting channels that are both highly weighted and consistently available maximises the proportion of subjects for which all inputs are present, avoiding reliance on masking and zero-padding at inference time. Availability-corrected weights are reported separately in the supplementary table 23 to provide insight into the model’s per-channel preferences independent of recording configuration. The availability corrected channels show a hierarchy of central *>* frontal *>* parietal *>* occipital *>* temporal. For these corrected weightings frontal channels (Fz, F4, F3) show a higher weighting across all channels, and for REM and wake the frontal channels are on par with central channels (apart from the dominant Cz).

Comparing performance between the clinical SleepWearables (SW) control group and the external MASS controls (table 22), we observed notable variability despite the absence of reported sleep disorders. This likely reflects differences in cohort clinical characteristics among the SW controls, the higher number of missing channels in the group, or simply the limited sample size available from each site (which constrains generalization across datasets). Additional EEG channels and multimodal configurations did not reduce the performance gap between SW and MASS; we see slight reductions in the performance gap for N1 and N3 using the 4 EEG + EOG model, compared to single EEG. However, the inclusion of EOG markedly reduced the inter-dataset gap in REM, suggesting that EOG improve cross-cohort generalization for REM classification.

Despite these gains, REM classification in RBD remained below thresholds typical of human inter-scorer agreement for REM in healthy or sleep-disordered breathing cohorts (*κ ≈* 0.69 [29]; 90.5% agreement in [20]). It should be noted that no comparable inter-rater benchmark exists specifically for REM in RBD. The results presented in this study may not exhibit clinically acceptable performance, but they show that integrating the full PSG into a sleep staging model improves performance in a mixed clinical cohort, and may reduce the number of PSGs required to obtain robust performance. By selecting the most important channels for staging, performance further improved. Future studies should be focused on developing models using all available inputs in real-life clinical settings, prior to identifying the most informative channels and minimising the sensor input.

Complex models (particularly those which include transformer blocks, such as AttnSleep) require large amounts of data to outperform classical models. This is particularly true when the task is more challenging, for example, the data more heterogenous. These findings should be interpreted in light of several constraints. The EEG channel weights are normalised to sum to one, and performance metrics are averaged across cross-validation folds. Moreover, fusion is performed after feature extraction, with modalities combined prior to temporal modelling. More integrated fusion approaches that retain richer modality-specific information prior to prediction may yield improved results. A further limitation concerns the structure of the clinical cohort. The SleepWearables data were collected across three sites with differing equipment, sampling rates, and recruitment patterns. Differences observed between RBD and non-RBD subjects may therefore be partially confounded by site or age effects rather than reflecting RBD pathology alone. With only 22 RBD subjects in the retrained cohort, the sample size precluded formal multivariate adjustment for site and age, or site-stratified subgroup analyses with adequate statistical power. We mitigated site-related amplitude variability through per-subject z-score normalisation and harmonised preprocessing across datasets, but residual site and demographic confounding cannot be excluded. Larger, more demographically balanced multi-site cohorts will be needed to disentangle disease-specific effects from acquisition and population effects.

## 10 Conclusions

This study evaluated a classical feature-based Random Forest model and a state-of-the-art deep learning model for automatic sleep staging in a mixed clinical cohort including RBD patients. When applied out-of-the-box to the clinical SW data, the Random Forest model outperformed AttnSleep, with the largest difference in REM classification in RBD subjects. Training on the clinical SW+MASS dataset and incorporating multimodal inputs notably improved the performance of AttnSleep, although these gains did not extend to the RBD subgroup. Guided by the learned modality weights, adding EOG to a reduced four-channel EEG model matched the full multimodal configuration overall while achieving the best performance in RBD subjects, indicating that the composition of the sensor set matters more than its size. These findings highlight the importance of adapting models to clinical populations and identifying physiologically relevant signals before prioritising minimal sensor set-ups. Athletes, particularly retired contact sport athletes, represent a high-priority group, as cumulative head impacts are associated with increased risk and progression to neurodegenerative disease. More broadly, RBD is a well-established prodromal marker of *α*-synucleinopathies, with a large proportion of patients eventually developing conditions such as Parkinson’s disease or dementia with Lewy bodies. However, detection remains limited due to reliance on resource-intensive PSG and under-recognition of symptoms, underscoring the need for automated, scalable approaches based on physiological sleep signals. Future systems should emphasize adaptive, multimodal architectures trained on diverse, clinically representative datasets.

## 11 Data availability

Access to the Oxford Parkinson’s Disease Centre (OPDC) SleepWearables PSG dataset used in this study is available through a formal submission to the Data Access Committee who will review this and either support, decline, or request further information. See https://www.dpag.ox.ac.uk/opdc/research/external-collaborations for more information.

## 12 Code availability

The code used in this work is publicly available at on GitHub (https://github.com/katarinamg/multi-modal-sleep-staging-).

## 13 Acknowledgements

The authors extend their sincere gratitude to all the participants who took part in this study and made this research possible. They are grateful to Oxford Biomedical Research Centre (BRC) and Parkinson’s UK for funding this project. They also thank the research hospital staff who assisted with in clinic data collection.

## Supplementary material

### A Random forest features

**Table 17:** Features extracted from EEG, EOG and EMG signals used as input to the random forest sleep staging model.

| Channel | Category | Name | Description |
| --- | --- | --- | --- |
| EEG | Time | Zero Crossing Rate | Number of times the EEG crosses its mean reference line. [43] |
| EEG | Time | Hjorth parameters | Activity, mobility and complexity computed from variances of derivatives [44] |
| EEG | Time | Time-domain properties | Low-power and log-amplitude of derivatives up to the 10th derivative, computed per mini-epoch. |
| EEG | Time | Percent differential | 75th minus 25th percentile of amplitude in the mini-epoch. [45] |
| EEG | Time | Mean / Min / Max coastline | Coastline = sum of absolute derivatives per mini-epoch; mean, min and max computed for each 30 s epoch. [46] |
| EEG | Frequency | STFT magnitude | Short-time Fourier transform magnitude computed per clinically relevant band. |
| EEG | Frequency | Relative spectral power | Relative power ratios for $\delta$, $\theta$, $\alpha$, $\beta$, $\gamma$ bands. ($RSP_{total}$ = sum of band RSPs). [43] |
| EEG | Frequency | Spectral edge frequency | $SEF_{95}$: frequency below which 95% of power is contained; often higher in REM. [43] |
| EEG | Non-linear | TKEO | Mean and SD of Teager–Kaiser Energy Operator for $\delta$, lower-α (8–10 Hz) and $\alpha$. [45] |
| EEG | Non-linear | Square-energy operator | Mean and SD of the square-energy operator per clinically relevant band. [47] |
| EOG | Time | Autocorrelation peak | Peak of autocorrelation per mini-epoch; enhances REM-related patterns. [46] |
| EOG | Time | Variance | Variance computed per mini-epoch. |
| EOG | Time | Max peak | Maximum absolute peak (positive or negative) in the mini-epoch. [46] |
| EOG | Time | Second max peak | Second-largest absolute peak in the mini-epoch. [46]) |
| EOG | Time | Differential & max differential | Average differential per mini-epoch; report average and maximum across 30 s. [19] |
| EOG | Frequency | Power ratio (0–4 Hz) | Power in 0–4 Hz band divided by total spectral power. [43] |
| EOG | Wavelet | DWT (Haar / DB2) | 4-level DWT; use max amplitude of level-4 inverse DWT as feature — larger values indicate REM. [46] |
| EOG | Non-linear | Permutation entropy | Permutation entropy (order 10) per mini-epoch; measures complexity. [45] |
| EEG & EOG | Time | Cross-correlation p-value | Pearson correlation p-value testing relationship between EEG and EOG. [19] |
| EEG & EOG | Frequency | Magnitude-squared coherence | Measure of synchrony between EEG and EOG signals. [43] |
| EMG | Time | Atonia index | Percent of amplitude values $\leq 1 \mu V$ (excluding 1–2 $\mu V$), averaged per 30 s epoch. [48]) |
| EMG | Time | Energy | Mean rectified amplitude per epoch. [49] |
| EMG | Time | 75th percentile | Value below which 75% of amplitude values lie. [50] |
| EMG | Time | Entropy | Variability computed from histogram (256 bins). [50] |
| EMG | Time | Motor activity | Algorithm-derived movement measure and threshold; used to obtain movement duration. [51] |
| EMG | Frequency | Fractal exponent | Negative slope of log–log spectral density. [43] |
| EMG | Frequency | Absolute gamma power | Mean power in 30–100 Hz band. [43] |
| EMG | Frequency | Relative power | Power in 12.5–21 Hz divided by total in 8–32 Hz band. [50] |
| EMG | Frequency | Spectral edge frequency | Frequency below which 95% of total power is located. [50] |
| NA | Time | Hours recorded | Hours into PSG recording (progression through night) used as a feature. ([19]) |

### B Convolutional architectures per modality

**Table 18:** EEG branch (per EEG channel) — multi-resolution CNN (MRCNN).

| Branch | Layer | Kernel | Stride | Padding | Notes |
| --- | --- | --- | --- | --- | --- |
| Fine | Conv1d(1→64) | 50 | 6 | 24 | BN, GELU |
|  | MaxPool1d | 8 | 2 | 4 |  |
| | Dropout | — | — | — | $p=0.5$ |
|  | Conv1d(64→128) | 8 | 1 | 4 | BN, GELU |
|  | Conv1d(128→128) | 8 | 1 | 4 | BN, GELU |
|  | MaxPool1d | 4 | 4 | 2 |  |
| Coarse | Conv1d(1→64) | 400 | 50 | 200 | BN, GELU |
|  | MaxPool1d | 4 | 2 | 2 |  |
| | Dropout | — | — | — | $p=0.5$ |
|  | Conv1d(64→128) | 7 | 1 | 3 | BN, GELU |
|  | Conv1d(128→128) | 7 | 1 | 3 | BN, GELU |
|  | MaxPool1d | 2 | 2 | 1 |  |
| Concat(time) → Dropout → SEBasicBlock($128 \rightarrow C_{\text{feat}}$) → AdaptiveAvgPool1d($T_{\text{out}}$) | | | | | |

**Table 19:**
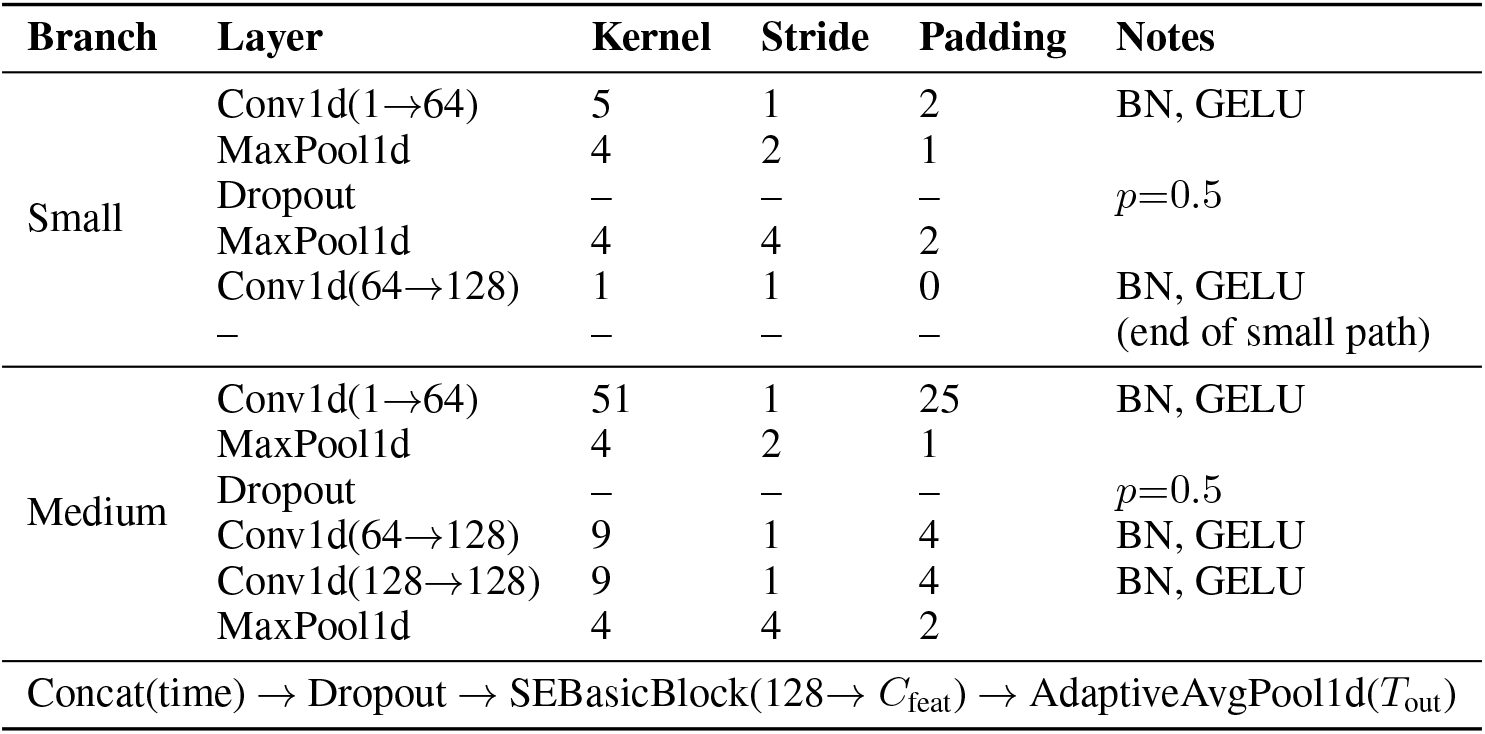
ECG branch.

| Branch | Layer | Kernel | Stride | Padding | Notes |
| --- | --- | --- | --- | --- | --- |
| Small | Conv1d(1→64) | 5 | 1 | 2 | BN, GELU |
|  | MaxPool1d | 4 | 2 | 1 |  |
| | Dropout | — | — | — | $p=0.5$ |
|  | MaxPool1d | 4 | 4 | 2 |  |
|  | Conv1d(64→128) | 1 | 1 | 0 | BN, GELU |
|  | — | — | — | — | (end of small path) |
| Medium | Conv1d(1→64) | 51 | 1 | 25 | BN, GELU |
|  | MaxPool1d | 4 | 2 | 1 |  |
| | Dropout | — | — | — | $p=0.5$ |
|  | Conv1d(64→128) | 9 | 1 | 4 | BN, GELU |
|  | Conv1d(128→128) | 9 | 1 | 4 | BN, GELU |
|  | MaxPool1d | 4 | 4 | 2 |  |
| Concat(time) → Dropout → SEBasicBlock($128 \rightarrow C_{\text{feat}}$) → AdaptiveAvgPool1d($T_{\text{out}}$) | | | | | |

**Table 20:**
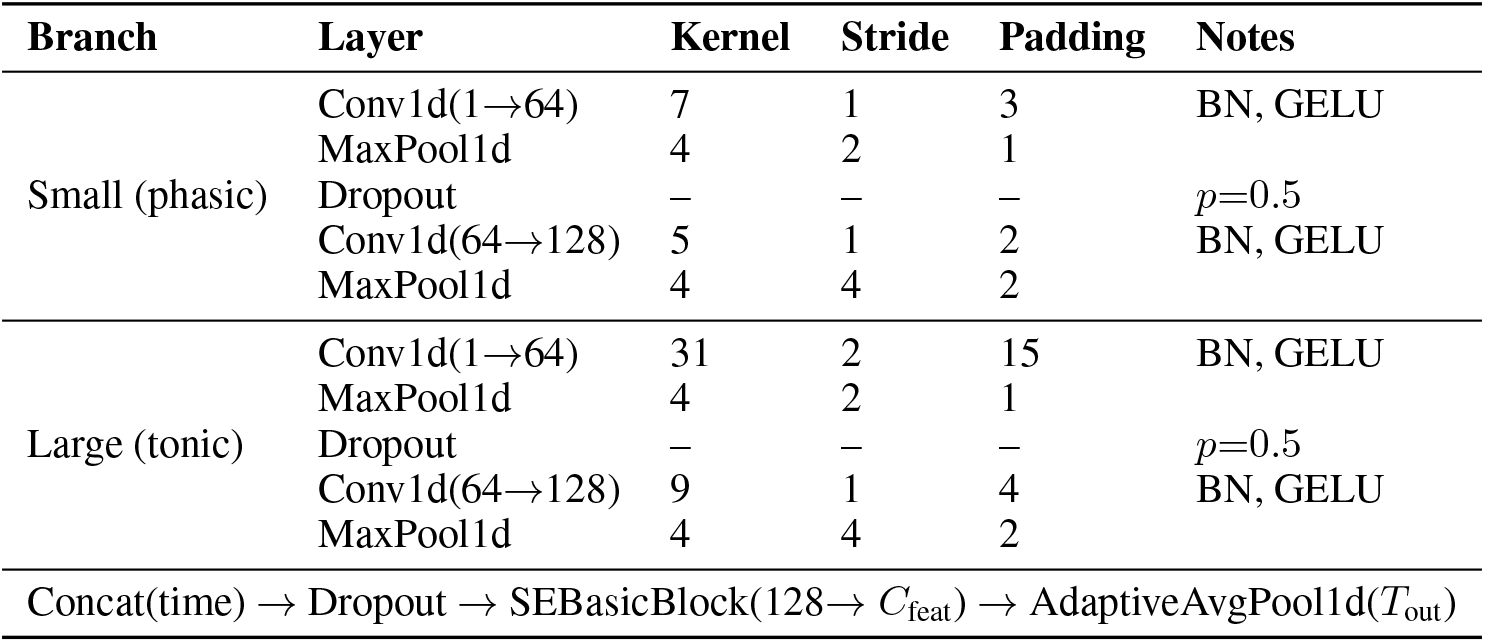
EMG branch.

**Table 21:** EOG branch.

| Branch | Layer | Kernel | Stride | Padding | Notes |
| --- | --- | --- | --- | --- | --- |
| Small | Conv1d(1→64) | 51 | 1 | 25 | BN, GELU |
|  | MaxPool1d | 4 | 2 | 1 |  |
| | Dropout | – | – | – | $p=0.5$ |
|  | Conv1d(64→128) | 9 | 1 | 4 | BN, GELU |
|  | MaxPool1d | 4 | 4 | 2 |  |
| Large | Conv1d(1→64) | $K_{\text{large}}=251$ | 1 | 125 | BN, GELU |
|  | MaxPool1d | 4 | 2 | 1 |  |
| | Dropout | – | – | – | $p=0.5$ |
|  | Conv1d(64→128) | 9 | 1 | 4 | BN, GELU |
|  | MaxPool1d | 4 | 4 | 2 |  |
| Concat(time) → Dropout → SEBasicBlock($128 \rightarrow C_{\text{feat}}$) → AdaptiveAvgPool1d($T_{\text{out}}$) | | | | | |

### C Dataset-Specific Performance

**Figure 5:**
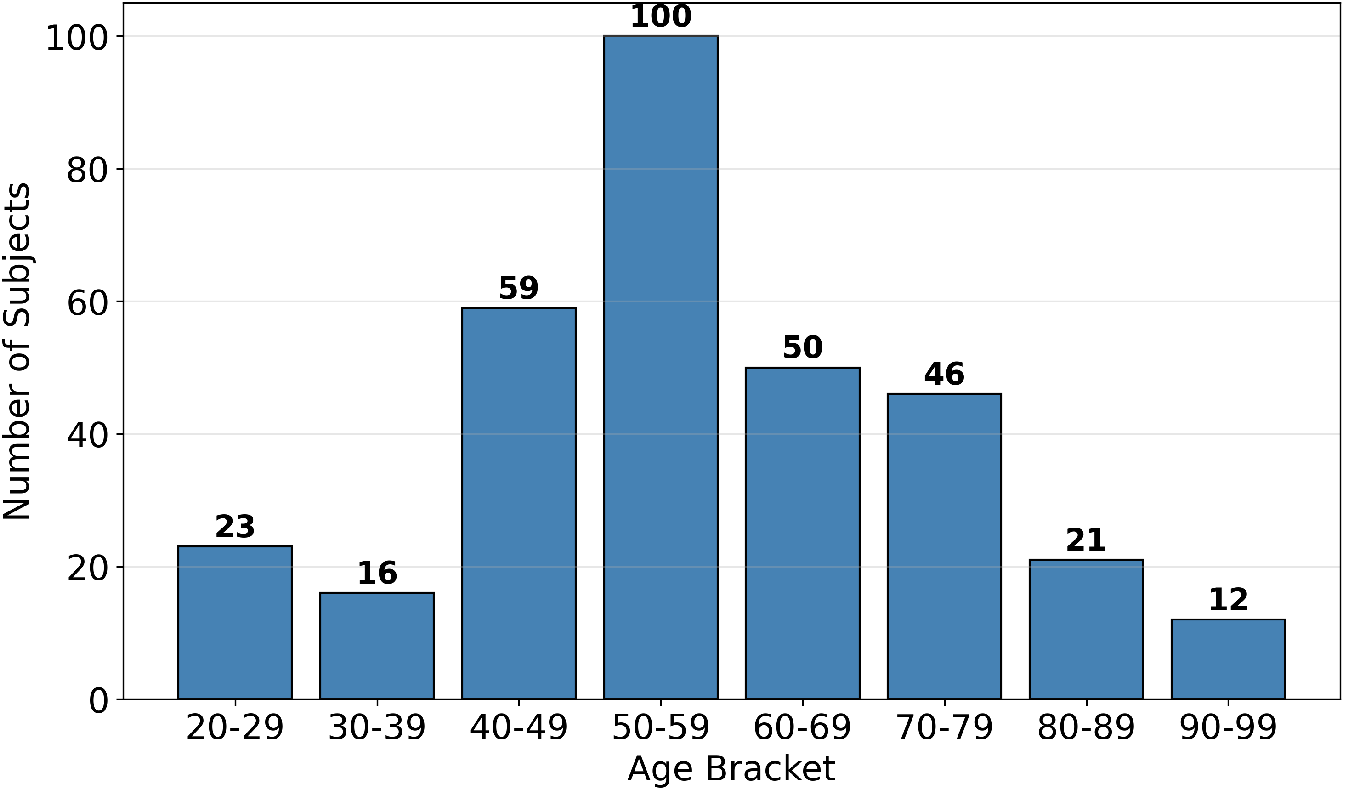
Distribution of subjects across age brackets in the combined SHHS and Sleep-EDF-78 dataset.

**Table 22:** Stage-level Cohen’s *κ* for Healthy Controls in SW and MASS datasets across model configurations.

| Model | Dataset | Wake | N1 | N2 | N3 | REM |
| --- | --- | --- | --- | --- | --- | --- |
| 1-EEG | SW | 0.79 | 0.20 | 0.55 | 0.77 | 0.59 |
|  | MASS | 0.88 | 0.43 | 0.70 | 0.59 | 0.71 |
| Multimodal | SW | 0.78 | 0.22 | 0.46 | 0.76 | 0.63 |
|  | MASS | 0.88 | 0.49 | 0.75 | 0.66 | 0.75 |
| 4-EEG | SW | 0.80 | 0.24 | 0.53 | 0.75 | 0.62 |
|  | MASS | 0.88 | 0.48 | 0.71 | 0.58 | 0.74 |
| 4-EEG + EOG | SW | 0.81 | 0.28 | 0.57 | 0.77 | 0.69 |
|  | MASS | 0.88 | 0.48 | 0.71 | 0.59 | 0.75 |

### D Modality presence rate and corrected importance

**Table 23:**
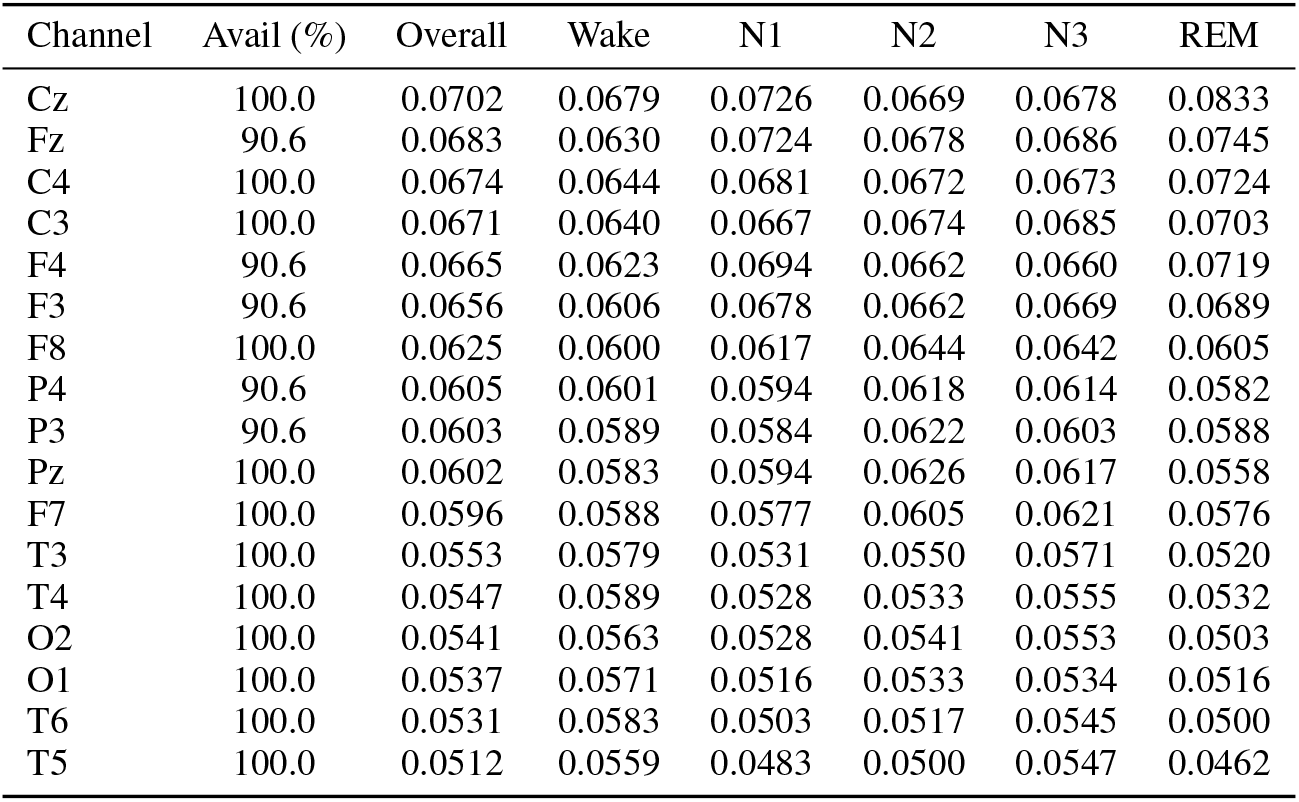
EEG channel availability and corrected weights (All subjects).

| Channel | Avail (%) | Overall | Wake | N1 | N2 | N3 | REM |
| --- | --- | --- | --- | --- | --- | --- | --- |
| Cz | 100.0 | 0.0702 | 0.0679 | 0.0726 | 0.0669 | 0.0678 | 0.0833 |
| Fz | 90.6 | 0.0683 | 0.0630 | 0.0724 | 0.0678 | 0.0686 | 0.0745 |
| C4 | 100.0 | 0.0674 | 0.0644 | 0.0681 | 0.0672 | 0.0673 | 0.0724 |
| C3 | 100.0 | 0.0671 | 0.0640 | 0.0667 | 0.0674 | 0.0685 | 0.0703 |
| F4 | 90.6 | 0.0665 | 0.0623 | 0.0694 | 0.0662 | 0.0660 | 0.0719 |
| F3 | 90.6 | 0.0656 | 0.0606 | 0.0678 | 0.0662 | 0.0669 | 0.0689 |
| F8 | 100.0 | 0.0625 | 0.0600 | 0.0617 | 0.0644 | 0.0642 | 0.0605 |
| P4 | 90.6 | 0.0605 | 0.0601 | 0.0594 | 0.0618 | 0.0614 | 0.0582 |
| P3 | 90.6 | 0.0603 | 0.0589 | 0.0584 | 0.0622 | 0.0603 | 0.0588 |
| Pz | 100.0 | 0.0602 | 0.0583 | 0.0594 | 0.0626 | 0.0617 | 0.0558 |
| F7 | 100.0 | 0.0596 | 0.0588 | 0.0577 | 0.0605 | 0.0621 | 0.0576 |
| T3 | 100.0 | 0.0553 | 0.0579 | 0.0531 | 0.0550 | 0.0571 | 0.0520 |
| T4 | 100.0 | 0.0547 | 0.0589 | 0.0528 | 0.0533 | 0.0555 | 0.0532 |
| O2 | 100.0 | 0.0541 | 0.0563 | 0.0528 | 0.0541 | 0.0553 | 0.0503 |
| O1 | 100.0 | 0.0537 | 0.0571 | 0.0516 | 0.0533 | 0.0534 | 0.0516 |
| T6 | 100.0 | 0.0531 | 0.0583 | 0.0503 | 0.0517 | 0.0545 | 0.0500 |
| T5 | 100.0 | 0.0512 | 0.0559 | 0.0483 | 0.0500 | 0.0547 | 0.0462 |

**Table 24:** Corrected modality weights across sleep stages (All subjects).

| Modality | All_Avail (%) | Overall | Wake | N1 | N2 | N3 | REM |
| --- | --- | --- | --- | --- | --- | --- | --- |
| EEG | 100.0 | 0.312 | 0.377 | 0.301 | 0.290 | 0.299 | 0.287 |
| ECG | 98.0 | 0.249 | 0.247 | 0.220 | 0.251 | 0.309 | 0.214 |
| EOG | 98.4 | 0.199 | 0.194 | 0.225 | 0.191 | 0.177 | 0.227 |
| EMG | 100.0 | 0.173 | 0.196 | 0.159 | 0.156 | 0.192 | 0.177 |

## Notes

### Competing Interest Statement

The authors have declared no competing interest.

### Author Declarations

The South Central Oxford Research Ethics Committee gave ethical approval for this work, with reference number 17/SC/0631.

### Summary of Updates

This version was revised due to an error in the preprocessing pipeline of certain datasets used in the work (resulting in lower performance of the models). Furthermore, one subject that was diagnosed as a control, has been found to have RBD. The changes do not significantly change the overall outcome of the results, but the results and figures (from Table 6 and onwards) have been updated accordingly.

## References

[1] Michael J Sateia. International classification of sleep disorders-third edition. Chest, 146(5):1387–1394, 2014.

[2] Marc Baltzan, Christine Yao, Daniel Rizzo, et al. Dream enactment behavior: review for the clinician. Journal of Clinical Sleep Medicine, 16(11):1949–1969, 2020.

[3] Seung-Gul Kang, In-Young Yoon, Sang-Ah Lee, Jong-Hyun Han, and Tae-Ho Kim. Rem sleep behavior disorder in the general population: prevalence and clinical characteristics. Sleep, 36(8):1147–1152, 2013.

[4] José Haba-Rubio, Birgit Frauscher, Pedro Marques-Vidal, Jérémy Toriel, Nicolas Tobback, David Andries, Martin Preisig, Peter Vollenweider, and Raphael Heinzer. Prevalence and determinants of rem sleep behavior disorder in the general population. Sleep, 41(2):zsx197, 2018.

[5] Ronald B Postuma, Alex Iranzo, Michele Hu, Birgit Högl, Bradley F Boeve, Raffaele Manni, Wolfgang H Oertel, Isabelle Arnulf, Luigi Ferini-Strambi, Monica Puligheddu, Elena Antelmi, Valerie Cochen De Cock, Dario Arnaldi, Brit Mollenhauer, Aleksandar Videnovic, Karel Sonka, Ki-Young Jung, Dieter Kunz, Yves Dauvilliers, Federica Provini, Simon J Lewis, Jitka Buskova, Milena Pavlova, Anna Heidbreder, Jacques Y Montplaisir, Joan Santamaria, Thomas R Barber, Ambra Stefani, Erik K St.Louis, Michele Terzaghi, Annette Janzen, Smandra Leu- Semenescu, Guiseppe Plazzi, Flavio Nobili, Friederike Sixel-Doering, Petr Dusek, Frederik Bes, Pietro Cortelli, Kaylena Ehgoetz Martens, Jean-Francois Gagnon, Carles Gaig, Marco Zucconi, Claudia Trenkwalder, Ziv Gan-Or, Christine Lo, Michal Rolinski, Philip Mahlknecht, Evi Holzknecht, Angel R Boeve, Luke N Teigen, Gianpaolo Toscano, Geert Mayer, Silvia Morbelli, Benjamin Dawson, and Amelie Pelletier. Risk and predictors of dementia and parkinsonism in idiopathic rem sleep behaviour disorder: a multicentre study. Brain, 142(3):744–759, 02 2019.

[6] John W. Adams, Victor E. Alvarez, Jesse Mez, Benjamin R. Huber, Yorghos Tripodis, Wen Xia, Guang Meng, Courtney A. Kubilus, Kelly Cormier, Patrick T. Kiernan, et al. Association of probable rem sleep behavior disorder with pathology and years of contact sports play in chronic traumatic encephalopathy. Acta Neuropathologica, 140(6):851–862, 2020.

[7] Maria Roig-Uribe, Marta Serradell, Ainhoa Muñoz-Lopetegi, Carles Gaig, and Alex Iranzo. Prior exposure to concussions in patients with isolated rem sleep behavior disorder. Sleep Medicine, 110:254–257, 2023.

[8] Alessia Collía, Alex Iranzo, Carles Gaig, Joan Santamaria, Marta Serradell, Ainhoa Muñoz-Lopetegi, et al. Former participation in professional football as an occupation in patients with isolated rem sleep behavior disorder leading to a synucleinopathy: a case–control study. Journal of Neurology, 270(7):3557–3564, 2023.

[9] NHS England. Referral to treatment (rtt) waiting times statistics – september 2025 statistical press no- tice. https://www.england.nhs.uk/statistics/statistical-work-areas/rtt-waiting-times/, Nov 2025. This RTT data release states the number of patient pathways waiting longer than 52 weeks (178,532) for treatment in September 2025. Accessed 2026.

[10] Raphael Vallat and Matthew P. Walker. A universal, open-source, high-performance tool for automated sleep staging. bioRxiv, 2021.

[11] Alexander Neergaard Olesen, Poul Jørgen Jennum, Emmanuel Mignot, and Helge Bjarup Dissing Sorensen. Automatic sleep stage classification with deep residual networks in a mixed-cohort setting. Sleep, 44(1):zsaa161, 2021.

[12] Huy Phan, Kaare Mikkelsen, Oliver Y. Chén, Philipp Koch, Alfred Mertins, and Maarten De Vos. Sleeptransformer: Automatic sleep staging with interpretability and uncertainty quantification. IEEE Transactions on Biomedical Engineering, 69(8):2456–2467, 2022.

[13] Hangyu Zhu, Wei Zhou, Cong Fu, Yonglin Wu, Ning Shen, Feng Shu, Huan Yu, Wei Chen, and Chen Chen. Masksleepnet: A cross-modality adaptation neural network for heterogeneous signals processing in sleep staging. IEEE Journal of Biomedical and Health Informatics, 27(5):2353–2364, 2023.

[14] Huijun Hu, Chen Zhuqi, and, et al. Research and application of deep learning-based sleep staging: Data, modeling, validation, and clinical practice. Data in Brief, S1087079224000017, 2024.

[15] B. Kemp, A.H. Zwinderman, B. Tuk, H.A.C. Kamphuisen, and J.J.L. Oberye. Analysis of a sleep-dependent neuronal feedback loop: the slow-wave microcontinuity of the eeg. IEEE Transactions on Biomedical Engineering, 47(9):1185–1194, 2000.

[16] Christian O’Reilly, Nadia Gosselin, and Julie Carrier. Montreal archive of sleep studies: an open-access resource for instrument benchmarking and exploratory research. Journal of sleep research, 23, 06 2014.

[17] Ary Goldberger, Luís Amaral, Leon Glass, Jeffrey Hausdorff, Plamen Ivanov, Roger Mark, Joseph Mietus, George Moody, Chung-Kang Peng, and H. Stanley. Physiobank, physiotoolkit, and physionet : Components of a new research resource for complex physiologic signals. Circulation, 101:E215–20, 07 2000.

[18] Stuart F. Quan, Barbara V. Howard, Conrad Iber, James P. Kiley, F. Javier Nieto, George T. O’Connor, David M. Rapoport, Susan Redline, John Robbins, Jonathan M. Samet, and ‡Patricia W. Wahl. The Sleep Heart Health Study: Design, Rationale, and Methods. Sleep, 20(12):1077–1085, 12 1997.

[19] Navin Cooray, Fernando Andreotti, Christine Lo, Mkael Symmonds, Michele T.M. Hu, and Maarten De Vos. Detection of rem sleep behaviour disorder by automated polysomnography analysis. Clinical Neurophysiology, 130(4):505–514, 2019.

[20] R. S. Rosenberg and S. Van Hout. The american academy of sleep medicine inter-scorer reliability program: Sleep stage scoring. Journal of Clinical Sleep Medicine, 9(1):81–87, 2013. Reported inter-scorer *κ* across large cohorts scored by multiple AASM-certified technologists.

[21] M. Younes and P. J. Hanly. Minimizing interrater variability in staging sleep by use of computer-derived features (mss). Sleep, 2016. Epoch-by-epoch agreement in 5-stage sleep scoring between two senior technologists was 78.9% (*κ ≈* 0.711).

[22] H. Danker-Hopfe, D. Kunz, G. Gruber, G. Klösch, J. L. Lorenzo, S. L. Himanen, B. Kemp, T. Penzel, J. Röschke, H. Dorn, A. Schlögl, E. Trenker, and G. Dorffner. Interrater reliability between scorers from eight european sleep laboratories in subjects with different sleep disorders. Journal of Sleep Research, 13(1):63–69, 2004. Reported overall Cohen’s *κ ≈* 0.68 across multiple disorders.

[23] Antoine Guillot and Valentin Thorey. Robustsleepnet: Transfer learning for automated sleep staging at scale. IEEE Transactions on Neural Systems and Rehabilitation Engineering, 29(1441–1451), 2021.

[24] Umaer Hanif, Anis Aloulou, Flynn Crosbie, Paul Bouchequet, Mounir Chennaoui, Thomas Andrillon, and Damien Leger. Deciphering insomnia: Benchmarking automated sleep staging algorithms for complex sleep disorders. Journal of Sleep Research, 2025.

[25] Emadeldeen Eldele, Zhenghua Chen, Chengyu Liu, Min Wu, Chee-Keong Kwoh, Xiaoli Li, and Cuntai Guan. An attention-based deep learning approach for sleep stage classification with single-channel eeg. IEEE Transactions on Neural Systems and Rehabilitation Engineering, 29:809–818, 2021.

[26] Gautam Krishna, Sameer Dharur, Oggi Rudovic, Pranay Dighe, Saurabh Adya, Ahmed Hussen Abdelaziz, and Ahmed H. Tewfik. Modality dropout for multimodal device directed speech detection using verbal and non-verbal features. CoRR, abs/2310.15261, 2023.

[27] Natalia Neverova, Christian Wolf, Graham W. Taylor, and Florian Nebout. Moddrop: adaptive multi-modal gesture recognition. IEEE Transactions on Pattern Analysis and Machine Intelligence, 37(10):2043–2056, 2015.

[28] J. Richard Landis and Gary G. Koch. The measurement of observer agreement for categorical data. Biometrics, 33(1):159–174, March 1977.

[29] Y. J. Lee, J. Y. Lee, J. H. Cho, and J. H. Choi. Interrater reliability of sleep stage scoring: A meta-analysis. Journal of Clinical Sleep Medicine, 18(1):193–202, January 2022.

[30] Mark I Boulos, Trevor Jairam, Tetyana Kendzerska, James Im, Anastasia Mekhael, and Brian J Murray. Normal polysomnography parameters in healthy adults: a systematic review and meta-analysis. The Lancet Respiratory Medicine, 7(6):533–543, 2019.

[31] Stanislas Chambon, Mathieu Galtier, Pierrick Arnal, Gilles Wainrib, and Alexandre Gramfort. A deep learning architecture for temporal sleep stage classification using multivariate and multimodal time series. IEEE Transactions on Neural Systems and Rehabilitation Engineering, 26(7):758–769, 2018.

[32] Alexander Neergaard Olesen, Poul Jørgen Jennum, Emmanuel Mignot, and Helge Bjarup Dissing Sorensen. Automatic sleep stage classification with deep residual networks in a mixed-cohort setting. Sleep, 44(1):z saa161, 2021.

[33] Yann LeCun, Yoshua Bengio, and Geoffrey Hinton. Deep learning. Nature, 521(7553):436–444, May 2015.

[34] Allan Rechtschaffen. A manual of standardized terminology, techniques and scoring system for sleep stages of human subjects. Ishiyaku Publishers, 1968.

[35] Conrad Iber. The aasm manual for the scoring of sleep and associated events: rules, terminology, and technical specification. American Academy of Sleep Medicine, 2007.

[36] Irit Berlad, Amichai Shlitner, Shaul Ben-Haim, and Peretz Lavie. Cardiac activity during sleep: autonomic correlates of the cyclic alternating pattern (cap). Sleep, 16(6):572–578, 1993.

[37] John Trinder, Jeremy Kleiman, Michael Carrington, Simon Smith, Simon Breen, Naresh Tan, and Yoon-Hee Kim. Autonomic activity during human sleep as a function of time and sleep stage. Journal of Sleep Research, 10(4):253–264, 2001.

[38] Thomas Andrillon, Yuval Nir, Richard J. Staba, Fabio Ferrarelli, Chiara Cirelli, Giulio Tononi, and Itzhak Fried. Sleep spindles in humans: insights from intracranial eeg and unit recordings. Journal of Neuroscience, 31(49):17821–17834, 2011.

[39] Ian M. Colrain. The k-complex: a 7-decade history. Sleep, 28(2):255–273, 2005.

[40] Sujith Vijayan, Kyle Q. Lepage, Nancy J. Kopell, and Sydney S. Cash. Frontal beta-theta network during rem sleep. eLife, 6:e18894, 2017.

[41] Scott Makeig and Martin Inlow. Lapses in alertness: coherence of fluctuations in performance and eeg spectra. Electroencephalography and Clinical Neurophysiology, 86(1):23–35, 1993.

[42] I. I. Goncharova, D. J. McFarland, T. M. Vaughan, and J. R. Wolpaw. Emg contamination of eeg: spectral and topographical characteristics. Clinical Neurophysiology, 114(9):1580–1593, 2003.

[43] Kristína Šušmáková and Anna Krakovská. Discrimination ability of individual measures used in sleep stages classification. Artificial Intelligence in Medicine, 44:261–277, 11 2008.

[44] Shayan Motamedi-Fakhr, Mohamed Moshrefi-Torbati, Martyn Hill, Catherine M. Hill, and Paul R. White. Signal processing techniques applied to human sleep eeg signals—a review. Biomedical Signal Processing and Control, 10:21–33, 3 2014.

[45] Tarek Lajnef, Sahbi Chaibi, Perrine Ruby, Pierre Emmanuel Aguera, Jean Baptiste Eichenlaub, Mounir Samet, Abdennaceur Kachouri, and Karim Jerbi. Learning machines and sleeping brains: Automatic sleep stage classification using decision-tree multi-class support vector machines. Journal of Neuroscience Methods, 250:94– 105, 7 2015.

[46] Benjamin D. Yetton, Mohammad Niknazar, Katherine A. Duggan, Elizabeth A. McDevitt, Lauren N. Whitehurst, Negin Sattari, and Sara C. Mednick. Automatic detection of rapid eye movements (rems): A machine learning approach. Journal of Neuroscience Methods, 259:72–82, 2 2016.

[47] Athanasios Tsanas, Max A. Little, Patrick E. McSharry, and Lorraine O. Ramig. Nonlinear speech analysis algorithms mapped to a standard metric achieve clinically useful quantification of average parkinson’s disease symptom severity. Journal of the Royal Society Interface, 8:842, 6 2010.

[48] Raffaele Ferri, Francesco Rundo, Mauro Manconi, Giuseppe Plazzi, Oliviero Bruni, Alessandro Oldani, Luigi Ferini-Strambi, and Marco Zucconi. Improved computation of the atonia index in normal controls and patients with rem sleep behavior disorder. Sleep Medicine, 11:947–949, 10 2010.

[49] Yu Liang Hsu, Ya Ting Yang, Jeen Shing Wang, and Chung Yao Hsu. Automatic sleep stage recurrent neural classifier using energy features of eeg signals. Neurocomputing, 104:105–114, 3 2013.

[50] S. Charbonnier, L. Zoubek, S. Lesecq, and F. Chapotot. Self-evaluated automatic classifier as a decision-support tool for sleep/wake staging. Computers in Biology and Medicine, 41:380–389, 6 2011.

[51] Rune Frandsen, Miki Nikolic, Marielle Zoetmulder, Lykke Kempfner, and Poul Jennum. Analysis of automated quantification of motor activity in rem sleep behaviour disorder. Journal of Sleep Research, 24:583–590, 10 2015.

